# Combining SNP-to-gene linking strategies to pinpoint disease genes and assess disease omnigenicity

**DOI:** 10.1101/2021.08.02.21261488

**Authors:** Steven Gazal, Omer Weissbrod, Farhad Hormozdiari, Kushal Dey, Joseph Nasser, Karthik Jagadeesh, Daniel Weiner, Huwenbo Shi, Charles Fulco, Luke O’Connor, Bogdan Pasaniuc, Jesse M. Engreitz, Alkes L. Price

## Abstract

Although genome-wide association studies (GWAS) have identified thousands of disease-associated common SNPs, these SNPs generally do not implicate the underlying target genes, as most disease SNPs are regulatory. Many SNP-to-gene (S2G) linking strategies have been developed to link regulatory SNPs to the genes that they regulate in *cis*, but it is unclear how these strategies should be applied in the context of interpreting common disease risk variants. We developed a framework for evaluating and combining different S2G strategies to optimize their informativeness for common disease risk, leveraging polygenic analyses of disease heritability to define and estimate their *precision* and *recall*. We applied our framework to GWAS summary statistics for 63 diseases and complex traits (average *N*=314K), evaluating 50 S2G strategies. Our optimal combined S2G strategy (cS2G) included 7 constituent S2G strategies (Exon, Promoter, 2 fine-mapped *cis*-eQTL strategies, EpiMap enhancer-gene linking, Activity-By-Contact (ABC), and Cicero), and achieved a precision of 0.75 and a recall of 0.33, more than doubling the precision and/or recall of any individual strategy; this implies that 33% of SNP-heritability can be linked to causal genes with 75% confidence. We applied cS2G to fine-mapping results for 49 UK Biobank diseases/traits to predict 7,111 causal SNP-gene-disease triplets (with S2G-derived functional interpretation) with high confidence. Finally, we applied cS2G to genome-wide fine-mapping results for these traits (not restricted to GWAS loci) to rank genes by the heritability linked to each gene, providing an empirical assessment of disease omnigenicity; averaging across traits, we determined that the top 200 (1%) of ranked genes explained roughly half of the heritability linked to all genes. Our results highlight the benefits of our cS2G strategy in providing functional interpretation of GWAS findings; we anticipate that precision and recall will increase further under our framework as improved functional assays lead to improved S2G strategies.

## Introduction

While genome-wide association studies (GWAS) have successfully identified thousands of disease-associated loci, they generally do not identify the underlying causal variants, target genes, cell-types and biological functions, thus limiting the translation of GWAS findings into discoveries that will enhance disease treatment^1–3^. Although recent advances in fine-mapping techniques have improved our ability to nominate causal variants^4–7^, identifying the underlying target genes remain a critical challenge, as causal variants are predominantly regulatory SNPs^8–11^ that do not necessarily regulate the closest genes^12–17^. Large gene expression quantitative trait loci (eQTL) data sets^18,19^ have proven useful in linking disease variants to their target genes through colocalization analyses^20–24^ or transcriptome-wide association studies^14,17,25^, but explain a limited proportion of disease heritability^15,26,27^, likely due to limited representation of disease-relevant cell-types/states^28^. Many other functional assays and computational approaches have recently been developed to link regulatory SNPs to their target genes in *cis* in a broad set of cell-types^26,29–38^; for example, EpiMap enhancers^37^ and ABC enhancers34,38 are linked to their target genes using correlation of enhancer activity with gene expression across cell-types and a quantitative combination of enhancer activity and 3D contact frequencies, respectively. Combining SNP-to-gene (S2G) linking strategies has previously been proposed as an appealing approach to improve SNP-to-gene linking^39–42^. However, it is currently unclear how S2G strategies should be prioritized in the context of GWAS, limiting our ability to pinpoint disease genes.

Here, we developed a framework for evaluating and combining S2G strategies to optimize their informativeness for human disease, leveraging polygenic analyses of disease heritability. In particular, we defined an S2G strategy’s *precision* and *recall* for identifying disease genes by evaluating the heritability enrichment of SNPs linked to a *critical gene set*, relying on the hypothesis that a precise S2G strategy should maximize the heritability linked to the critical gene set compared to the heritability linked to all genes. We applied this framework to GWAS summary statistics for 63 diseases and complex traits (average *N*=314K), evaluating 50 S2G strategies and constructing an optimal combined S2G strategy (cS2G) informed by GWAS data. We applied cS2G to fine-mapping results for 49 diseases and complex traits from the UK Biobank^7,43^ to pinpoint disease genes at thousands of GWAS loci. Finally, we applied cS2G to genome-wide fine-mapping results for these diseases/traits to rank genes by the heritability linked to each gene, enabling an empirical assessment of the “omnigenic model”^44–46^.

## Results

### Overview of Methods

A SNP-to-gene (S2G) linking strategy is defined as an assignment of linking scores between each SNP and zero or more candidate target genes (Figure 1a), such that each SNP has a sum of linking scores ≤1 (see Methods). The proportion of SNPs linked to one or more genes can vary widely, e.g. 2.3% for the exon strategy (which links SNPs located in exons ± 20bp) vs. 100% for the Closest TSS strategy (see below). All analyses were restricted to 19,995 genes (see Methods).

**Figure 1:**
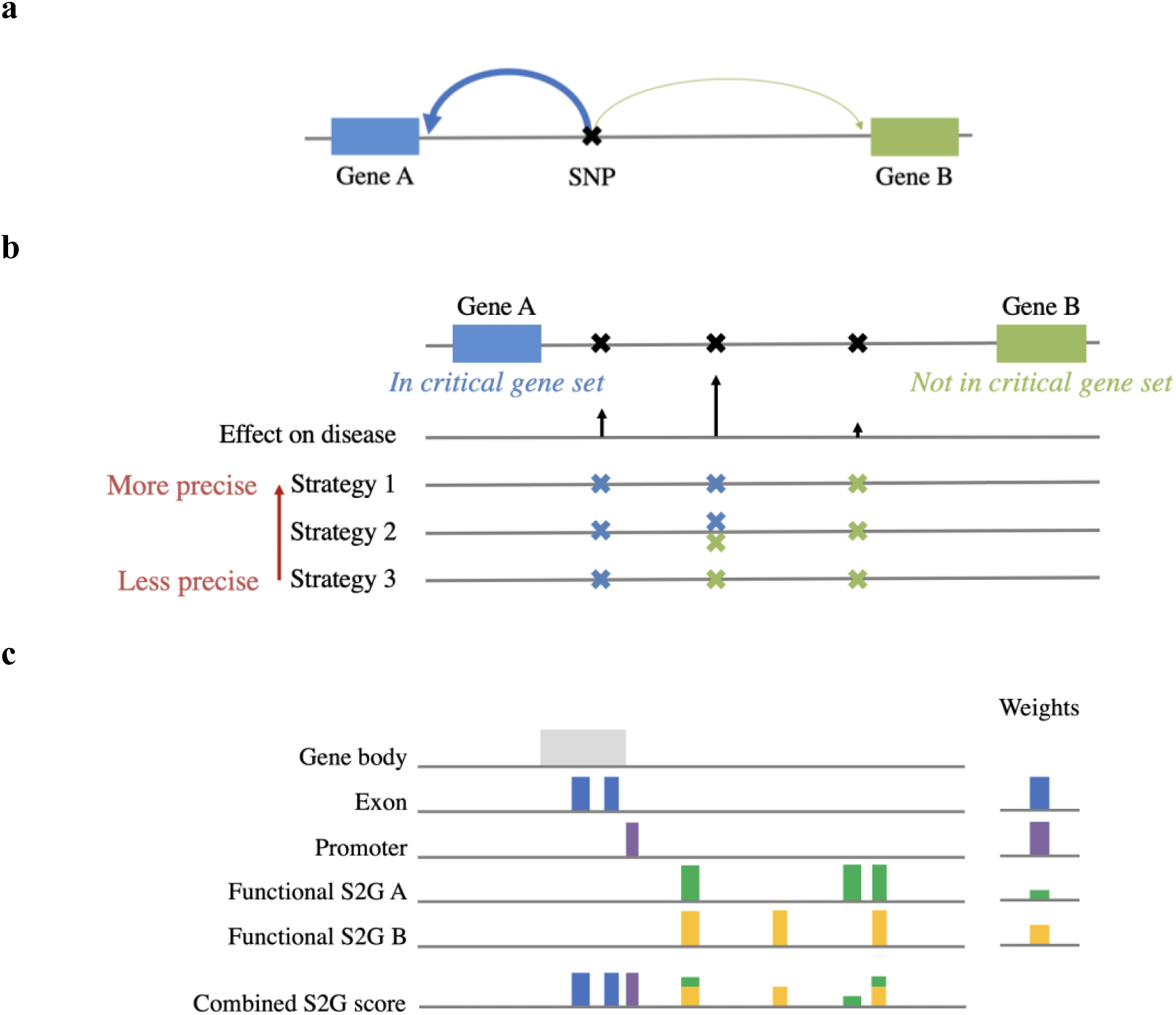
Overview of S2G framework. **(a)** Toy example of SNP linked to two genes (arrow widths denote linking scores). **(b)** Toy example of using critical gene sets to define precision. Strategy 1 (which links the middle SNP with high effect on disease to the gene from the critical gene set) is more precise than strategy 2 (which links the middle SNP to both genes), which is more precise than strategy 3 (which links the middle SNP to the gene that is not from the critical gene set). **(c)** Toy example of combined S2G strategy. The combined S2G strategy is a linear combination of constituent S2G strategies.

We analyzed 13 main S2G strategies that link SNPs to their target genes in *cis* (Table 1 and Methods): 2 high-confidence strategies (Exon^47^ and Promoter^11,30^), 3 non-functionally informed strategies (Gene body^48,47,49^, Gene±100kb^50^, and Closest TSS), 2 fine-mapped *cis*-eQTL^26^ strategies (using GTEx^18^ and eQTLGen^19^ datasets), 3 enhancer-gene linking strategies^29^ (using Roadmap29,30,32 and EpiMap^37^ enhancers informed by gene expression and Activity-By-Contact (ABC)^34,38^ enhancers informed by Hi-C), 2 Promoter capture Hi-C (PCHi-C)^31,35^ strategies, and 1 single-cell ATAC-seq (scATAC-seq) strategy (Cicero blood/basal^33,36^). Linking scores of fine-mapped GTEx *cis*-eQTL, and Roadmap, EpiMap, and ABC enhancer-gene linking strategies were defined by using the maximum across all available tissues and cell-types (as with ABC in refs.^38,51^). Correlations between the 13 main S2G strategies (Supplementary Table 1) indicate low concordance between the strategies (average correlation of 0.05 between the 10 functionally informed S2G strategies). We analyzed 50 S2G strategies in total (Supplementary Table 2), including the 13 main strategies, 26 additional strategies based on physical distance to TSS, 4 main strategies restricted to blood and immune tissues and cell-types (for comparison purposes), 3 strategies based on all *cis*-eQTL (instead of fine-mapped *cis*-eQTL), 2 strategies based on Hi-C intensity, and 2 combined strategies (GeneHancer^39^ and Open Targets^42^). In our primary analyses, each S2G strategy was restricted to the gene(s) with the highest linking score, as we observed that this led to slightly higher precision. We note that our evaluation of these S2G strategies is impacted by their widely varying underlying biosample sizes (see Methods), in addition to differences in functional assays and SNP-to-gene linking methods.

**Table 1:** Description of the 13 main SNP-to-gene (S2G) strategies. For each of 13 main S2G strategies (in 6 categories), we provide a brief description and report the % SNPs linked (proportion of common SNPs that are linked to at least one gene) and *h*^*2*^ coverage (meta-analyzed across 63 independent traits). A description of all 50 S2G strategies analyzed is provided in Supplementary Table 2.

| S2G strategy | Description | % SNPs linked | $h^2$ coverage |
| --- | --- | --- | --- |
| **High confidence (2)** |  |  |  |
| Exon* | Exons<sup>47</sup> +/- 20bp | 2.3% | 9.8% |
| Promoter* | TSSs<sup>49</sup> +/- 1kb ∩ promoter annotations<sup>11,30</sup> | 2.0% | 5.8% |
| **Non-functionally informed (3)** |  |  |  |
| Gene body | Gene body<sup>48,47,49</sup> | 43.1% | 55.4% |
| Gene±100kb | Gene body<sup>48,47,49</sup> +/- 100kb<sup>50</sup> | 73.2% | 85.4% |
| Closest TSS | Gene with closest TSS<sup>49</sup> | 100% | 100% |
| **Fine-mapped *cis*-eQTL (2)** |  |  |  |
| GTEx fine-mapped *cis*-eQTL* | Fine-mapped GTEx v8 *cis*-eQTLs<sup>18,26</sup> in 54 cell-types | 9.5% | 19.4% |
| eQTLGen fine-mapped blood *cis*-eQTL* | Fine-mapped eQTLGen *cis*-eQTLs<sup>19,26</sup> in blood | 2.2% | 6.2% |
| **Enhancer-gene linking (3)** |  |  |  |
| Roadmap enhancer-gene linking | Correlation between Roadmap enhancers and gene expression across 127 cell-types<sup>29,30,32</sup> | 8.6% | 16.3% |
| EpiMap enhancer-gene linking* | Correlation between EpiMap enhancers and gene expression across 833 cell-types<sup>37</sup> | 7.5% | 13.8% |
| Activity-By-Contact (ABC)* | Hi-C linked enhancers in 167 cell-types<sup>34,38</sup> | 7.7% | 17.3% |
| **PCHi-C (2)** |  |  |  |
| Jung PCHi-C | Promoter capture Hi-C in 27 cell-types<sup>35</sup> | 39.9% | 43.9% |
| Javierre PCHi-C blood | Promoter capture Hi-C in 17 blood cell-types<sup>31</sup> | 27.2% | 35.5% |
| **scATAC-seq (1)** |  |  |  |
| Cicero blood/basal* | Correlation between scATAC-seq peaks and gene promoter peaks across 61,806 blood/basal cells<sup>33,36</sup> | 1.0% | 2.9% |

\* included in our combined S2G strategy (cS2G).

To evaluate each S2G strategy’s informativeness for pinpointing disease genes, we aimed to define and estimate parameters that correspond to an S2G strategy’s heritability coverage (proportion of total disease SNP-heritability (*h*^*2*^) that is linked to genes; *h*^*2*^ coverage), precision (proportion of disease *h*^*2*^ linked to genes that is linked to the correct target gene), and recall (proportion of total disease *h*^*2*^ that is linked to the correct target gene). First, we defined *h*^*2*^ *coverage* as the proportion of *h*^*2*^ explained by all SNPs linked to one or more genes (weighted by their linking scores). Second, we defined *precision* as the relative excess *h*^*2*^ enrichment of SNPs linked to a critical gene set (see below) vs. SNPs linked to all genes, as compared to the (gold-standard) Exon S2G strategy; this definition is based on the intuition that a precise S2G strategy is more likely (than an imprecise strategy) to link a disease SNP to a critical gene (Figure 1b). We note that this definition relies on the hypothesis that genes in the critical gene set enriched for causal disease genes (as observed empirically, see below; however, not all genes in the critical gene set have to be causal), and the hypothesis that the Exon S2G strategy is a perfectly precise strategy (even though it suffers from low *h*^*2*^ coverage). Third, we defined *recall* as the product of the *h*^*2*^ coverage and precision. We estimated these quantities using polygenic analyses of disease heritability by applying stratified LD score regression (S-LDSC) with the baseline-LD model (v2.2)^11,52,53^ to 63 independent diseases and complex traits (average *N* = 314K; Supplementary Table 3), meta-analyzing results across traits. We jointly analyzed SNP annotations derived from the 50 S2G strategies for ∼10M SNPs with a minor allele count ≥5 in a 1000 Genomes Project European reference panel^54^.

Our definitions of precision and recall rely on a critical gene set (see above). We used a non-trait-specific *training* critical gene set to construct an optimal combined S2G strategy (see below), and trait-specific *validation* critical gene sets to evaluate the optimal combined S2G strategy while avoiding overfitting (for comparison purposes, we also used the validation critical gene sets to evaluate individual S2G strategies). We defined the training critical gene set as the top 10% of genes with the most highly constrained exons and conserved promoters, and the validation critical gene set for a given trait as the top 10% of genes ranked by the PoPS method^51^; the excess overlap between the training and validation critical gene sets was limited (median (across traits) overlap of 20% of genes in each gene set, vs. 10% expected by chance). These gene sets attained high heritability enrichment (e.g. 10.8±0.8x and 14.1±0.7x for a SNP annotation constructed by restricting to exons of these gene sets, vs. 4.4±0.2x for a SNP annotation constructed by restricting to exons of all genes; Supplementary Table 4), validating their informativeness for disease.

We constructed combined S2G strategies as linear combinations of linking scores from constituent S2G strategies (Figure 1c). Specifically, for each SNP *j* and gene *g* we computed a combined S2G score

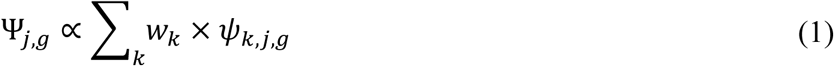

where *ψ*_*k,j,g*_ is the linking score between SNP *j* and gene *g* for S2G strategy *k*, and *w*_*k*_ is the weight associated to strategy *k*; the combined scores Ψ_*j,g*_ were subsequently normalized for each SNP *j* so that Σ_*g*_ Ψ_*j,g*_ ≤ 1 (we allowed Σ_*g*_ Ψ_*j,g*_ < 1 to allow for incomplete SNP-to-gene linking). To estimate the optimal weights *w*_*k*_, we developed an optimization framework to identify the weights maximizing the recall while constraining precision (defined using the training critical gene set) to be ≥0.75; the precision and recall of the resulting combined S2G strategies were subsequently evaluated using the validation critical gene sets.

Further details are provided in the Methods section. We have released open-source software implementing our framework (see Code Availability), and have made all S2G strategies, SNP annotations and critical gene sets analyzed publicly available (see Data Availability).

### Evaluation of S2G strategies

We estimated the *h*^*2*^ coverage, precision and recall for the 50 S2G strategies (Table 1 and Supplementary Table 2), meta-analyzed across the 63 independent diseases and complex traits (Supplementary Table 3); we used the (trait-specific) validation critical gene sets to perform these evaluations. Results for the 13 main S2G strategies are reported in Figure 2 and Supplementary Figure 1, and results for all 50 S2G strategies are reported in Supplementary Table 5. The Exon and Promoter strategies attained high precision (1.00 for Exon (by definition) and 0.80 for promoters), but low *h*^2^ coverage (0.06-0.10) and thus low recall (0.05-0.10). On the other hand, the Closest TSS strategy attained low precision (0.34), but the highest *h*^2^ coverage (1.00) and recall (0.34). In addition to Exon and Promoter, 5 other main S2G strategies attained high precision (>0.5) but low recall (0.02-0.13): the 2 fine-mapped *cis*-eQTL strategies, the 2 enhancer-gene linking strategies informed by gene expression, and the scATAC-seq strategy. Interestingly, S2G strategies using fine-mapped *cis*-eQTLs^26^ attained significantly higher precision than S2G strategies using all *cis*-eQTL (0.68 ± 0.07 vs. 0.40 ± 0.04 in GTeX, 0.81 ± 0.11 vs. 0.29 ± 0.03 in eQTLGen), consistent with previous reports of low precision for strategies using all *cis*-eQTL^17,55^ and emphasizing the advantage of fine-mapped *cis*-eQTL for more precise analyses of GWAS data.

**Figure 2:**
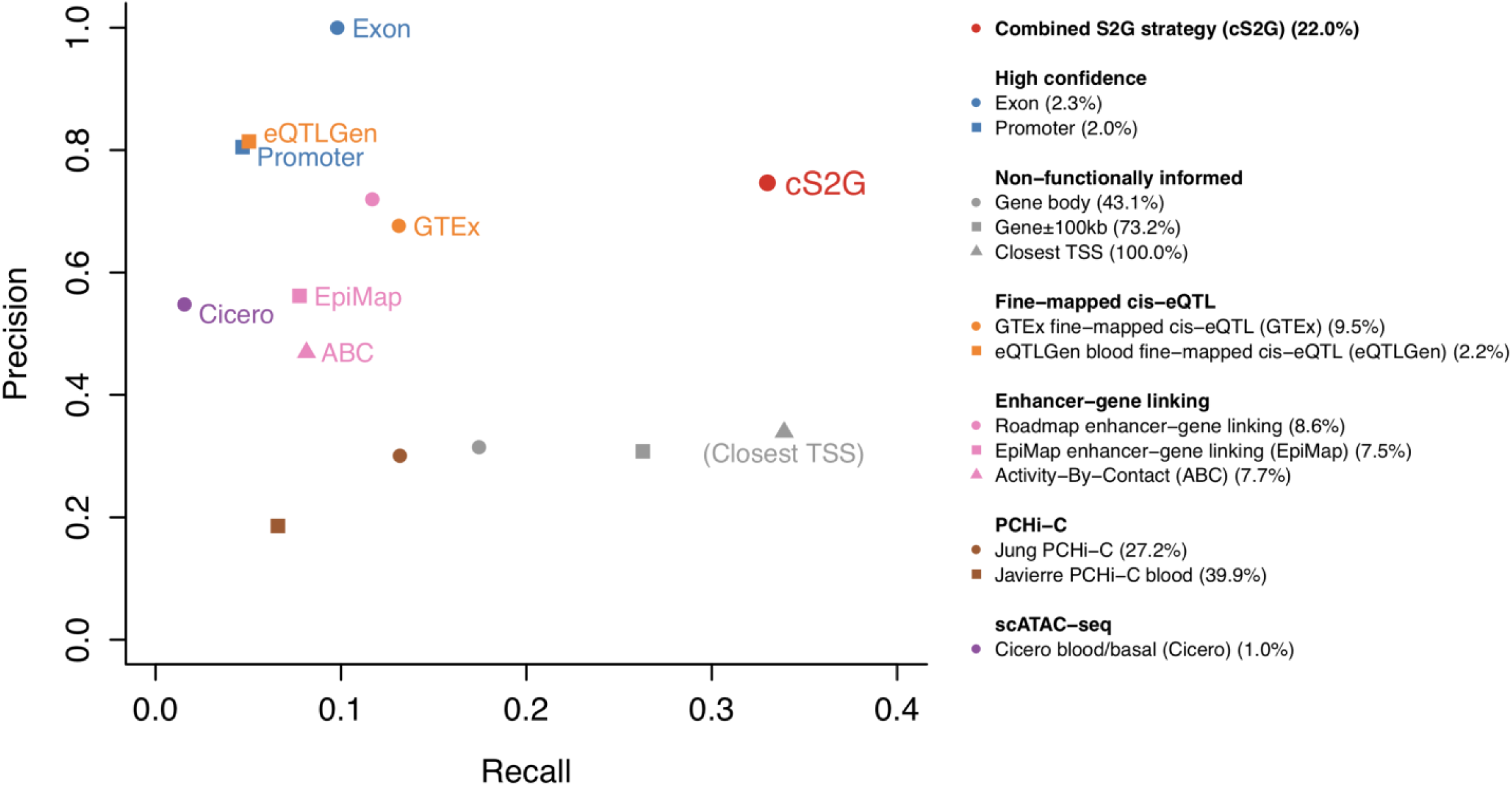
Accuracy of individual S2G strategies and combined S2G (cS2G) strategy. We report the precision and recall of the 13 main S2G strategies from **Table 1** and the cS2G strategy (estimated using trait-specific validation critical gene sets and meta-analyzed across 63 independent traits). Colored font denotes the cS2G strategy and its 7 constituent S2G strategies (gray font in parentheses denotes the Closest TSS strategy). Numbers in parentheses in legend denotes the proportion of common SNPs that are linked to at least one gene (as in **Table 1**). We note that our evaluation of these S2G strategies is impacted by their widely varying underlying biosample sizes (see Methods), in addition to differences in functional assays and SNP-to-gene linking methods. Standard errors are reported in Supplementary Figure 1, and numerical results are reported in Supplementary Table 5; standard errors for all S2G strategies linking >2.5% of common SNPs were ≤0.12 for precision and ≤0.03 for recall, with smaller standard errors for S2G strategies linking larger proportions of common SNPs.

The 50 S2G strategies included 27 S2G strategies based on physical distance to TSS: 7 S2G strategies defined by different ranges of distance to closest TSS (<1kb, 1-5kb, etc.; each SNP links to ≤1 gene), and 20 S2G strategies defined by different ranks of distance to TSS (closest TSS, 2nd closest TSS, etc.; each SNP links to 1 gene). Results for these S2G strategies are reported in Supplementary Figure 2 and Supplementary Table 5. As expected, we observed that proximal closest TSS are likely to implicate target genes (e.g. precision of 0.78 for closest TSS range <1kb), whereas distal closest TSS are much less likely to implicate target genes (e.g. precision of 0.15 for closest TSS range 100-500kb). We further determined that closest TSS are moderately likely to implicate target genes: precision of 0.34, decreasing to 0.17 for 2nd closest TSS and 0.062 for 5th closest TSS. The mean value of 0.043 for 6th-20th closest TSS suggests that genes located in the same regions as causal disease genes have a slightly elevated probability of being causal.

We next investigated whether functionally informed S2G strategies restricted to trait-specific tissues and cell-types were more precise for the corresponding traits. We compared our main S2G strategies defined using all available tissues and cell-types (GTEx fine-mapped *cis*-eQTL and Roadmap, EpiMap and ABC enhancer-gene linking) to corresponding strategies restricted to blood and immune tissues and cell-types, in analyses restricted to 11 autoimmune diseases and blood cell traits (average *N* = 257K; Supplementary Table 3). We determined that S2G strategies defined using all available tissues and cell-types achieved higher precision than S2G strategies restricted to blood and immune cell-types (Supplementary Figure 3), perhaps due to limited biosample size; these results support including all available cell-types in current efforts to pinpoint disease genes^17,27^ (see Discussion).

We report three lines of evidence to further validate our precision metric. First, we verified that the estimated precision of the Closest TSS strategy (0.34 ± 0.03) is consistent with previous studies (0.34 in ref.^14^, ∼0.50 in ref.^15^, 0.29 in ref.^16^, 0.35 in ref.^17^, and 0.27 ± 0.06 in ref.^56^), and that the estimated recall (*h*^*2*^ coverage times precision) of the GTEx fine-mapped *cis*-eQTL strategy (0.13 ± 0.01) is consistent with the proportion of *h*^2^ mediated by gene expression in GTEx tissues estimated using a different approach^27^ (0.11 ± 0.02). Second, we verified that estimates of precision (and hence recall) were similar when estimated using the (non-trait-specific) training critical gene set (used to optimize cS2G; see below) instead of the (trait-specific) validation critical gene sets (Supplementary Figure 4). Third, we verified that estimates of precision were similar for most S2G strategies when using an independent definition of precision (not relying on critical gene sets or polygenic analyses) based on two curated disease-associated lists of 577 linked sentinel SNP-gene pairs with the underlying genes validated with high confidence by Open Targets^42^ and 1,668 linked fine-mapped SNP-gene pairs validated using nearby fine-mapped protein-coding variants^51^ (see Methods and Supplementary Figure 5). Despite the overall concordance, we observed large differences in precision estimates for some S2G strategies (e.g. Closest TSS), as the curated SNP-gene pairs were preferentially ascertained for disease-associated SNPs in which the target gene was the closest gene: indeed, we observed an unusually high proportion of SNP-gene pairs involving genes with a small distance (< 10kb) to its closest TSS (57% and 67% for the two curated lists, vs. *h*^*2*^ coverage = 34% for the Closest TSS <10kb S2G strategy). Thus, we caution that curated disease-associated lists of linked SNP-gene pairs may be non-randomly ascertained, highlighting the potential benefits of polygenic analyses for evaluating S2G strategies.

**Figure 3:**
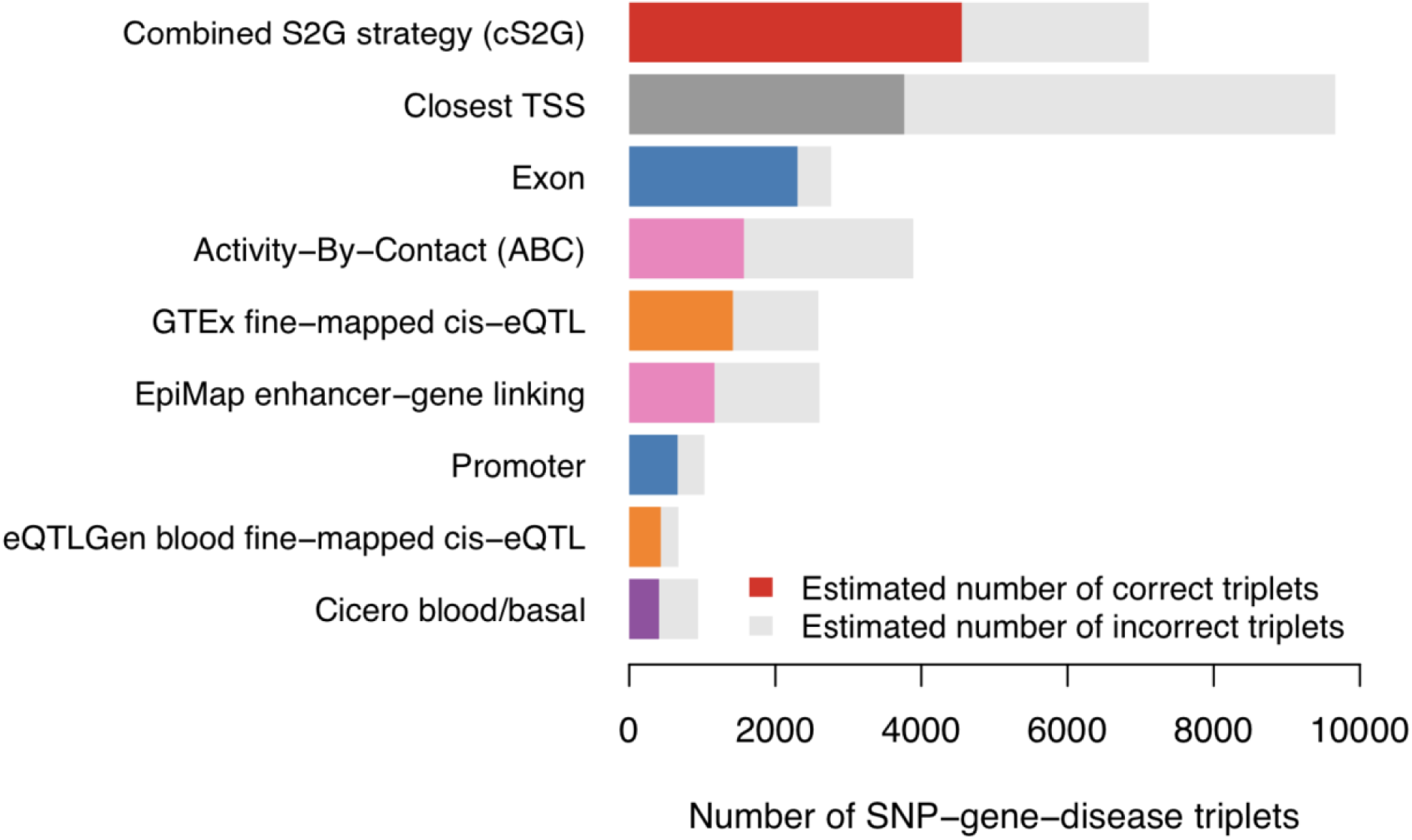
Number of SNP-gene-disease triplets identified by cS2G and other S2G strategies. We report the number of SNP-gene-disease triplets identified by cS2G, its 7 consistent strategies and Closest TSS S2G strategy. For each strategy, we estimated the number of correct triplets based on the mean confidence score across triplet; the estimated number of correct triplets is denoted as a colored bar, and the estimated number of incorrect triplets is denoted as a grey bar. The list of SNP-gene-disease triplets predicted by cS2G is reported in Supplementary Table 12. Numerical results, including numbers of SNP-gene-disease triplets identified by cS2G and all 13 main S2G strategies, and numbers of SNP-gene-disease triplets for the 13 main S2G strategies and cS2G, are reported in Supplementary Table 13.

**Figure 4:**
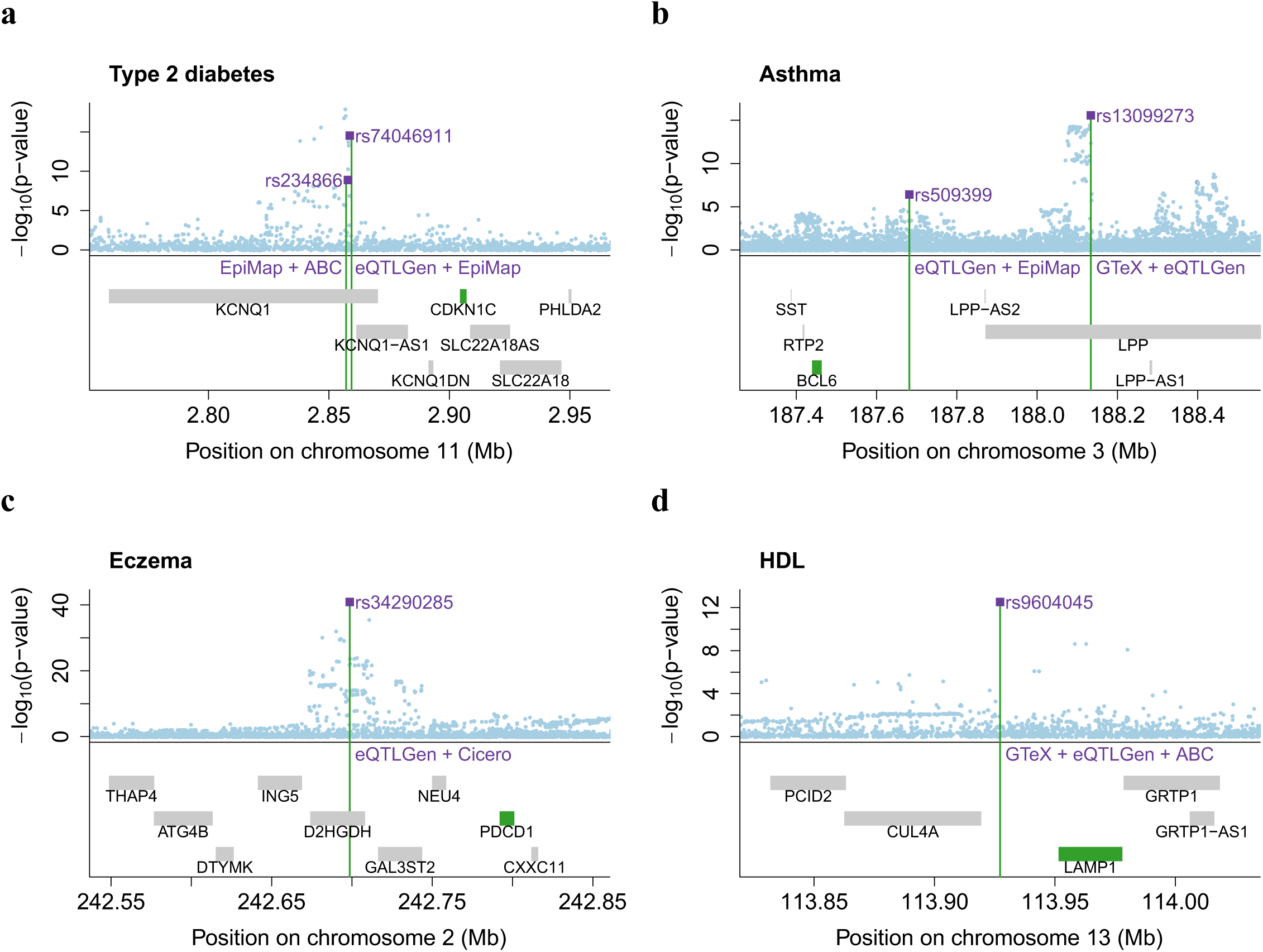
Examples of high-confidence SNP-gene-disease triplets identified by cS2G. We report four examples where cS2G predicts target genes for distal regulatory fine-mapped SNPs (i.e. not in promoter or gene body) for (**a**) type 2 diabetes, (**b**) asthma, (**c**) eczema, and (**d**) high-density lipoprotein (HDL) cholesterol. Fine-mapped SNPs are denoted as purple squares, target genes are denoted in green, and constituent S2G strategies implicating the target gene are denoted in purple. All fine-mapped SNPs in these examples have posterior inclusion probability (PIP) >0.9 for the corresponding disease/trait, except rs13099273 for asthma (PIP=0.58). S2G links for all 13 main S2G strategies are reported in Supplementary Table 12, and tissues/cell-types for constituent strategies of cS2G are reported in Supplementary Table 15. GTEx: GTEx fine-mapped *cis*-eQTL; eQTLGen: eQTLGen blood fine-mapped *cis*-eQTL; EpiMap: EpiMap enhancer-gene linking; ABC: Activity-By-Contact; Cicero: Cicero blood/basal.

**Figure 5:**
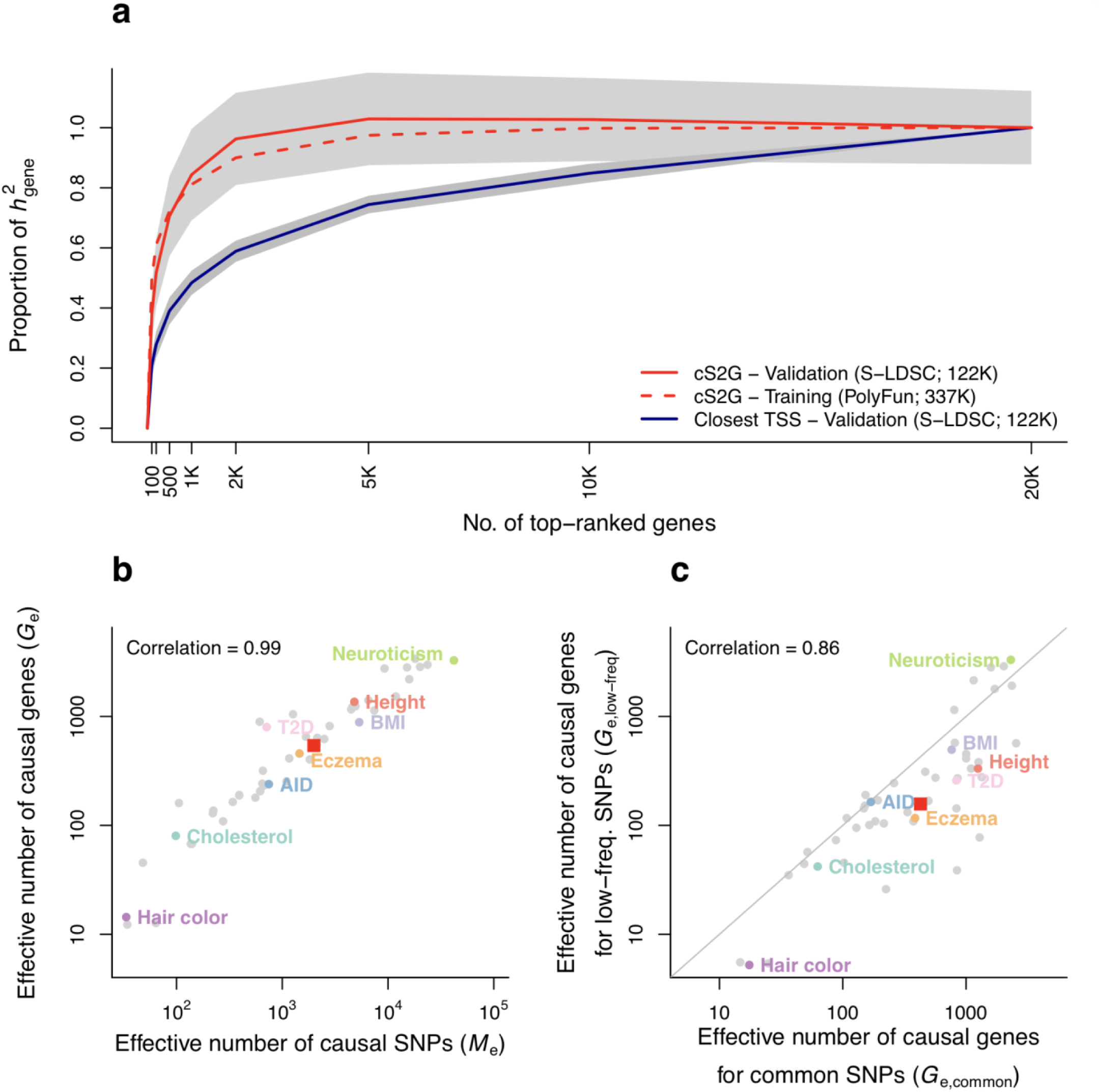
Empirical assessment of disease omnigenicity using cS2G. (**a**) We report the proportion of heritability linked to genes (*h*^2^_*gene*_) explained by genes ranked by top per-gene *h*^*2*^, as inferred using three approaches (see text). Grey shading denotes 95% confidence intervals for cS2G-validation and Closest TSS-validation. Results were meta-analyzed across 16 independent UK Biobank traits. (**b**) We report the effective number of causal SNPs^73^ (*M*_*e*_) and the effective number of causal genes (*G*_*e*_) for 49 UK Biobank diseases/traits, with representative traits in colored font. (**c**) We report the effective number of causal genes for per-gene heritabilities linked to common SNPs (*G*_*e,common*_) and the effective number of causal genes for per-gene heritabilities linked to low-frequency SNPs (*G*_*e,low-frea*_) for 49 UK Biobank diseases/traits, with representative traits in colored font. In (**b**) and (**c**), red squares denote median values across 16 independent traits and correlations are computed on log-scale values. Numerical results are reported in Supplementary Table 19 and Supplementary Table 21. AID: Autoimmune disease; BMI: Body mass index; Cholesterol: Total cholesterol; T2D: Type 2 diabetes.

In summary, we developed and validated a polygenic framework for evaluating S2G strategies, inferring high precision but low recall for many functionally informed S2G strategies, and low precision but relatively high recall for non-functionally informed strategies (such as Closest TSS).

### Combining S2G strategies

We constructed an optimal combined S2G strategy (cS2G) by considering linear combinations of linking scores from 10 functionally informed main S2G strategies (Table 1), maximizing the recall while constraining precision to be ≥0.75 using the (non-trait-specific) training critical gene set; we did not include the 3 non-functionally informed main S2G strategies (Table 1), because a fundamental goal of cS2G is to provide functional interpretation of GWAS findings. The resulting cS2G strategy included 7 constituent S2G strategies: Exon, Promoter, eQTLGen blood fine-mapped *cis*-eQTL, GTEx fine-mapped *cis*-eQTL, EpiMap enhancer-gene linking, ABC, and Cicero blood/basal (ordered from highest to lowest weight; Supplementary Table 6). The cS2G strategy linked 22% of common SNPs (minor allele frequency (MAF) ≥ 5%) to at least one gene and 99.6% of genes to at least one common SNP (average of 1.20 genes per linked common SNP and 79 common SNPs per linked gene); despite the high weights for Exon and Promoter, 43% of linked common SNPs were not linked to the gene with closest TSS. The number of common SNPs linked per gene was correlated to gene-body length (*r*^*2*^ = 0.13), but uncorrelated to gene-body length after correcting for exon and promoter length (*r*^*2*^ = 0.01). Strikingly, only 18% (resp. 3.3%) of the cS2G links were supported by at least 2 (resp. 3) of 7 constituent S2G strategies (Supplementary Table 7), consistent with the low correlations between the constituent S2G strategies (Supplementary Table 1). This provides a strong motivation for combining multiple S2G strategies.

We evaluated the cS2G strategy using the (trait-specific) validation critical gene sets and meta-analyzed the results across the 63 independent diseases and complex traits. The cS2G strategy attained *h*^2^ coverage of 0.44 (s.e. 0.01), precision of 0.75 (s.e. 0.06), and recall of 0.33 (s.e. 0.03) (Figure 2 and Supplementary Table 5), more than doubling the precision and/or recall of any individual strategy; this implies that 33% of SNP-heritability can be linked to causal genes with 75% confidence using cS2G. Notably, cS2G attained much higher precision than two previously proposed combined strategies, GeneHancer^39^ (0.14) and Open Targets^42^ (0.33) (Supplementary Table 5). We note that the precision of 0.75 for the cS2G strategy computed using the validation critical gene sets is independent of the threshold of precision ≥0.75 in the training critical gene set used to optimize the cS2G strategy (estimated precision using the training critical gene set was equal to 0.81 during the optimization process; Supplementary Table 8).

We performed 5 secondary analyses. First, we constructed a new S2G strategy in which we linked all the SNPs linked by cS2G (22% of common SNPs) to the gene with closest TSS. This strategy attained only slightly lower precision and recall than the cS2G strategy (0.70 vs. 0.75 and 0.31 vs. 0.33, respectively), indicating that the most important difference between the cS2G and Closest TSS strategies derives from the set of SNPs linked to genes, rather than the linking strategy applied to those SNPs (Supplementary Table 8); evaluation of S2G strategies defined by linking other sets of functional SNPs to the gene with closest TSS is of potential interest. Second, we expanded the set of S2G strategies provided as input when constructing the combined S2G strategy by adding the 3 non-functionally informed main S2G strategies from Table 1 (Gene body, Gene±100kb and Closest TSS), thus including all 13 main S2G strategies. The resulting combined strategy was identical to our primary cS2G combined strategy, indicating that Closest TSS provides no additional information. Third, we expanded the set of S2G strategies to include all 50 S2G strategies (Supplementary Table 2), not just the 13 main S2G strategies. The resulting combined strategy included 8 S2G strategies (Supplementary Table 9), attained higher recall (0.39, vs. 0.33 for cS2G), but lower precision (0.60 vs. 0.75). (We note that the estimated precision of this combined strategy using the training critical gene set was equal to 0.84 during the optimization process (Supplementary Table 8); the difference between 0.84 and 0.60 can be attributed to higher precision for some constituent S2G strategies in the training critical gene set compared to the validation critical gene sets (Supplementary Figure 4 and Supplementary Table 5). For this reason (and because a fundamental goal of cS2G is to provide functional interpretation of GWAS findings), we recommend the use of the cS2G strategy. Fourth, we constructed a new combined strategy by optimizing the *F1 score* (the harmonic mean of precision and recall^57^) instead of optimizing the recall while constraining precision to be ≥0.75. Once again, this strategy greatly outperformed the individual strategies (now with respect to the F1 score) in the validation critical gene sets (Supplementary Table 10), confirming that our framework is robust to the choice of optimization metric. Finally, to assess whether training and validating cS2G using different critical gene sets (but the same 63 independent traits) avoids overfitting, we randomly split the set of 63 independent traits in two, and performed training using one set of traits (using the training critical gene set) and validation using the other set of traits (using the validation critical gene sets). This procedure lead to combined S2G strategies with high precision (≥0.76, vs. 0.75 for cS2G in the analysis of 63 traits) and recall (≥0.30, vs. 0.33) (Supplementary Table 11), confirming that our primary cS2G combined strategy avoids overfitting.

In summary, we constructed an optimal combined S2G strategy (cS2G), incorporating 7 constituent S2G strategies, which more than doubles the precision and/or recall of any individual strategy; our evaluation of cS2G was based on validation critical gene sets that were distinct from the training critical gene set used to optimize cS2G.

### Leveraging the combined S2G strategy to pinpoint disease genes

We leveraged the high precision and recall of our combined S2G (cS2G) strategy (estimated at 0.75 and 0.33, respectively; see above) to predict target genes of candidate causal variants. We analyzed 9,670 predicted causal SNP-disease pairs with posterior inclusion probability (PIP) >0.50 from functionally informed fine-mapping of 49 UK Biobank diseases/traits^7,43^ (7,675 unique SNPs) (Supplementary Table 12).

Restricting to SNPs that had a linked gene with cS2G linking score >0.5 (average cS2G linking score = 0.95), we predicted 7,111 causal SNP-gene-disease triplets (5,384 unique SNPs; 3,401 unique genes) (Figure 3 and Supplementary Table 12). We note that the proportion of fine-mapped SNPs linked by the cS2G strategy (5,384/7,675 = 0.70) is higher than its *h*^2^ coverage (0.44) due to the excess of exon and promoter SNPs (2,629/5,384 SNPs are located in exons or promoters, consistent with the use of functional priors in fine-mapping analyses). The mean PIP of the 7,111 SNP-gene-disease triplets was equal to 0.80. We further assigned a confidence score to each of the triplets by multiplying the PIP of the constituent SNP and its precision (accounting for the excess of exon and promoter SNPs; see Methods, Supplementary Table 12 and Supplementary Table 13). The average confidence score was 0.64, implying that 64% of the 7,111 SNP-gene-disease triplets (4,554 triplets) predict the correct causal SNP and target gene (Figure 3). This number of correct triplets is 1.98 times larger than what could be attained using any individual functionally informed constituent S2G strategy, and 1.21 times higher than what could be attained using the Closest TSS strategy (despite 1.36 times fewer total triplets) (Figure 3 and Supplementary Table 13). In many instances, multiple causal SNPs were linked to the same gene (e.g. 119 genes were each linked to at least 5 different fine-mapped SNPs for 5 different diseases/traits; Supplementary Table 14), implying that a single gene can be causal for different diseases/traits using different causal SNP-gene links.

The SNP-gene-disease triplets predicted by cS2G included 2,163 triplets involving distal regulatory fine-mapped SNPs that were not in the gene body (or promoter) of the target gene, of which 532 were supported by at least 2 of the functionally informed constituent S2G strategies used by cS2G (Supplementary Table 12). We highlight 4 examples (Figure 4). First, for type 2 diabetes, two SNPs, rs234866 and rs74046911 (*r*^2^ = 0.02 (but *D*’ = 1)), both located in an intron of *KCNQ1* (initially reported as a candidate target gene^58,59^), were fine-mapped (PIP = 0.97 and 0.92, respectively) and both linked by cS2G to *CDKN1C* (third closest TSS; cS2G linking scores = 1.00 and 0.96, respectively) (Figure 4a). *CDKN1C* is a gene expressed in pancreas for which a rare coding mutation was previously linked to type 2 diabetes^60^, and has been nominated as a candidate target gene at this locus using methylation data^61^ and CRISPR-Cas9 genome editing^62^. *CDKN1C* was implicated by 3 of the functionally informed S2G strategies used by cS2G, including EpiMap enhancer-gene linking in endocrine pancreas (for both SNPs), identified by LDSC-SEG^50^ as a critical tissue for type 2 diabetes (see Supplementary Table 15). Second, for asthma, two independent SNPs, rs509399 and rs13099273 (*r*^2^ < 0.01), were fine-mapped (PIP = 0.96 and 0.58, resp.) and linked by cS2G to the target gene *BCL6* (third and sixth closest TSS, resp.; cS2G linking scores = 0.96 and 1.00, resp.) (Figure 4b). *BCL6* modulates the response of interleukin 4, known to be involved in asthma^63^, and has been linked to asthma in mice^64^ and humans^65^, but to our knowledge *BCL6* has not previously been implicated as an asthma gene using GWAS data. *BCL6* was implicated by 3 of the functionally informed S2G strategies used by cS2G, including EpiMap enhancer-gene linking in common myeloid progenitor CD34+ cells (for rs509399 only), identified by LDSC-SEG^50^ as a critical cell-type for asthma (*P* = 1.2 × 10^−4^; see Methods; also see ref.^66^). Third, for eczema, the SNP rs34290285 was fine-mapped (PIP = 0.99) and linked by cS2G to *PDCD1* (seventh closest TSS; cS2G linking score = 0.73) (Figure 4c). *PDCD1* is an immune-inhibitory receptor expressed in activated T cells, known to be implicated in Eczema. *PDCD1* has previously be linked to skin cancer^67^ and autoimmune diseases^68^, but to our knowledge *PDCD1* has not previously been implicated as an eczema gene using GWAS data. *PDCD1* was implicated by the 2 blood-informed S2G strategies used by cS2G (eQTLGen fine-mapped blood *cis*-eQTL and Cicero blood/basal). Two others functionally informed S2G strategies (GTEx fine-mapped *cis*-eQTL and EpiMap) linked rs34290285 to different genes (*GAL3ST2* and *D2HGDH*, respectively) with lower cS2G linking scores (0.21 and 0.05, respectively), highlighting the benefits of aggregating evidence from multiple S2G strategies to infer biological mechanisms. Fourth, for HDL cholesterol, the SNP rs9604045 was fine-mapped (PIP = 0.99) and linked by cS2G to *LAMP1* (closest TSS, although *CUL4A* is the closest gene; cS2G linking score = 0.97) (Figure 4d). While deficiency of lysosome associated membrane proteins (*LAMP1* and *LAMP2*) has been connected to cholesterol accumulation in mice^69,70^, to our knowledge *LAMP1* has not previously been implicated as an HDL gene using GWAS data. *LAMP1* was implicated by 3 of the functionally informed S2G strategies used by cS2G (GTEx fine-mapped *cis*-eQTL, eQTLGen fine-mapped blood *cis*-eQTL, and ABC). However, none of the S2G strategies implicating *LAMP1* involved a plausible critical tissue/cell-type for HDL cholesterol (e.g. liver, identified by LDSC-SEG^50^; Supplementary Table 15), despite the availability of S2G links for liver in GTeX and ABC. This result highlights both the benefit of aggregating S2G links across multiple cell-types to infer SNP-gene pairs, and the challenge of identifying the causal cell-type of action.

We extended our analyses to 222,842 potentially causal SNP-disease pairs with PIP>0.05 (instead of PIP>0.50) from functionally informed fine-mapping of 49 UK Biobank diseases/traits^7,43^. Restricting to SNPs that had a linked gene with cS2G linking score >0.5, we predicted 138,716 potentially causal SNP-gene-disease triplets (99,847 unique SNPs, 15,820 unique genes) (see Data Availability). We also analyzed 170,346 SNP-disease pairs from the NHGRI-EBI GWAS catalog^2^ (4,688 diseases/traits), with the caveat that these SNPs were not fine-mapped and have only a small probability of being causal (see below). Restricting to SNPs that had a linked gene with cS2G linking score >0.5, we predicted 78,499 potentially causal SNP-gene-disease triplets (49,313 unique SNPs, 13,349 unique genes) (see Data Availability).

Finally, to more broadly assess the application of cS2G to pinpoint disease genes using associated SNPs instead of fine-mapped SNPs, we analyzed a curated disease-associated list of 577 linked sentinel SNP-gene pairs (for ∼300 diseases and complex traits, partially distinct from the set of 63 traits used to construct the cS2G strategy) with the underlying genes validated with high confidence by Open Targets^42^ (see above). 356 of the 577 SNPs had a linked gene with cS2G linking score >0.5. The cS2G prediction of the target gene matched the Open Targets prediction for 205 of these 356 SNPs (58%); these included SNP-gene links between the high-density lipoprotein (HDL) cholesterol sentinel SNP rs4983559 (ref.^71^) with *AKT1* (second closest TSS), and the depression sentinel SNP rs915057 (located in an intron of *SYN2*; ref. ^72^) with *ESR2* (second closest TSS) (Supplementary Table 16). We observed that the discrepancy between 58% and our estimated precision (0.75) can be explained by the fact that Open Targets reports sentinel SNPs (rather than causal fine-mapped SNPs), which may be linked to different genes than causal SNPs, as we verified in analyses of UK Biobank traits (Supplementary Table 17). We thus recommend to apply our cS2G strategy to confidently fine-mapped SNPs in preference to sentinel SNPs.

In summary, our cS2G strategy validated 205 previously curated disease-associated SNP-gene pairs^42^, and predicted 7,111 causal SNP-gene-disease triplets with high confidence; these triplets represent, to our knowledge, the largest high-confidence SNP-gene-disease resource with S2G-derived functional interpretation to date.

### Leveraging the combined S2G strategy to empirically assess disease omnigenicity

Previous work has proposed an “omnigenic model” in which all genes expressed in disease-critical cell-types impact the function of core disease genes and thus impact disease heritability^44–46^. This work raises intense interest in estimating components of disease heritability contributed by each gene, but this has yet to be empirically assessed, due to the challenges of linking SNPs to genes. We leveraged our combined S2G (cS2G) strategy to estimate the heritability causally explained by SNPs linked to each gene for 49 UK Biobank diseases/traits (Supplementary Table 18) with functionally informed genome-wide fine-mapping results (not restricted to GWAS loci) available for all MAF ≥ 0.1% SNPs^7^. We partitioned genes into 8 gene sets ranked by per-gene heritability explained (top 100, 200, 500, etc ; see *x*-axis of Figure 5a) and re-estimated the heritability linked to each ranked gene set by running S-LDSC with the baseline-LD model^11,52,53^ on summary statistics computed from *N*=122K European-ancestry UK Biobank validation samples that were distinct from the *N*=337K British UK Biobank training samples used for fine-mapping (to avoid winner’s curse; see Methods and ref.^7^). We also estimated the effective number of causal genes (*G*_*e*_) for each trait (using fourth moments of per-gene effects, analogous to *M*_*e*_ for SNPs^73^), and estimated this quantity separately for per-gene heritabilities linked to common (MAF≥5%) and low-frequency (0.1%≤MAF<5%) SNPs.

The top 200 (resp. top 2,000) genes explained 52±6% (resp. 96±8%) of the disease heritability linked to genes in *cis* using the cS2G strategy (*h*^2^_*gene*_, which captures 53±3% of *h*^2^), meta-analyzed across a subset of 16 independent diseases/traits (Figure 5a and Supplementary Table 19) (we note that the estimate of *h*^2^_*gene*_/*h*^2^, previously termed *h*^2^ coverage, was higher than our previous estimate of 0.44 across 63 independent diseases/traits, due to the different set of diseases/traits analyzed). These estimates of proportions of *h*^2^_*gene*_ (based on *N*=122K new validation samples) are robust to winner’s curse, but estimates directly based on the *N*=337K training samples used for fine-mapping were very similar, implying minimal effects of winner’s curse (Figure 5a). The estimates of proportions of *h*^2^_*gene*_ explained by the top 200 genes varied widely across diseases/traits, from 25±5% (neuroticism) to 40±4% (height) to 86±18% (total cholesterol) (Supplementary Table 20; also see *G*_*e*_ estimates below). Results were similar when restricting to genes expressed in disease-critical cell-types^74^ (as proposed in ref. ^44^) (Supplementary Figure 6). Interestingly, repeating the analysis using the Closest TSS S2G strategy implicated a far more polygenic gene-level architecture that required the top 1,000 (resp. top 10,000) genes to explain 48±2% (resp. 85±2%) of *h*^2^_*gene*_ (Figure 5a and Supplementary Table 19); these results demonstrate the benefits of using more precise S2G strategies (e.g. 0.75 for cS2G vs. 0.34 for Closest TSS; Figure 2) to infer more accurately infer gene-level architectures. We caution that the primary analysis using cS2G may still slightly overestimate gene-level polygenicity, because even the cS2G strategy is not perfectly precise. We further caution that all of our findings pertain to effects of SNPs in *cis* (see Discussion), potentially leading to underestimation of gene-level polygenicity.

We estimated the effective number of causal genes (*G*_*e*_) for each trait. These estimates relied on per-gene heritabilities explained by causal SNPs, and were thus directly based on the *N*=337K samples used for fine-mapping; as noted above, the impact of winner’s curse on our analyses was minimal. Estimates of *G*_*e*_ varied widely, from 3,289 (neuroticism) to 1,375 (height) to 80 (total cholesterol) with a median of 540 (across 16 independent traits), and were strongly correlated (log-scale *r* = 0.99) to estimates of the effective number of independently associated SNPs (*M*_*e*_; median = 1,991), a SNP-based measure of disease/trait polygenicity^73^ (Figure 5b and Supplementary Table 21; the strong correlation provides a validation of both *G*_*e*_ and *M*_*e*_.).

We further estimated *G*_*e,common*_ (resp. *G*_*e,low-frea*_) by restricting per-gene heritabilities explained by causal SNPs to the heritability causally explained by common (resp. low-frequency) SNPs linked to each gene. Gene-level architectures were more polygenic for common vs. low-frequency SNPs, with median *G*_*e,common*_ of 427 vs. median *G*_*e,low-freq*_ of 157 (median ratio of 2.8) across the 16 independent traits (Figure 5c and Supplementary Table 21), consistent with more polygenic SNP architectures for common vs. low-frequency SNPs due to the action of negative selection (*M*_*e,common*_ / *M*_*e,low-freq*_ = 3.9 in ref. ^73^). Surprisingly, there was low concordance between genes underlying gene-level architectures for common vs. low-frequency SNPs, with a median (across the 16 independent traits) of only 26 shared genes among the top 200 genes ranked by their contribution to common and low-frequency variant heritability linked to genes (*h*^2^_*gene,common*_ and *h*^2^_*gene,low-freq*_, respectively), and a median correlation of only 0.02 between per-gene contributions to *h*^2^_*gene,common*_ and *h*^2^_*gene,low-freq*_ (Supplementary Figure 7 and Supplementary Table 21). Across all 49 diseases/traits, we identified only 13 unique genes that were included in the top 3 genes contributing to both *h*^2^_*gene,common*_ and *h*^2^_*gene,low-freq*_ for the same disease/trait (Supplementary Table 22), including *CDKN1C* for type 2 diabetes (further validating *CDKN1C* as a causal gene at this locus). Interestingly, we observed consistent excess overlap between the top 200 genes contributing to *h*^2^_*gene,common*_ (resp. *h*^2^_*gene,low-freq*_) and two disease-specific gene sets, Mendelian disorder genes^75^ (Supplementary Figure 8) and differentially expressed gene sets^50^ (Supplementary Figure 9), suggesting that common and low-frequency variant gene-level architectures are driven by different genes pertaining to similar biological processes.

In summary, our cS2G strategy provided a quantitative assessment of the “omnigenic model”^44–46^, implicating a far less polygenic gene-level architecture than the Closest TSS S2G strategy. We inferred a more polygenic gene-level architecture for common variants as compared to low-frequency variants, with little overlap between genes underlying these gene-level architectures despite shared biological processes.

## Discussion

We developed a polygenic framework to evaluate and combine S2G strategies, and validated our framework using two curated disease-associated lists of SNP-gene pairs^42,51^ (Supplementary Figure 5). We applied our framework to construct a combined S2G (cS2G) strategy that achieved a precision of 0.75 and a recall of 0.33, more than doubling the precision and/or recall of any individual strategy. We applied cS2G to fine-mapping results for 49 UK Biobank diseases/traits to predict 7,111 causal SNP-gene-disease triplets (with S2G-derived functional interpretation) with high confidence, including 2,163 triplets involving distal regulatory fine-mapped SNPs that were not in the gene body (or promoter) of the target gene; notable examples included *CDKN1C* in type 2 diabetes, *BCL6* in asthma, *PDCD1* in eczema, and *LAMP1* in HDL, all of which were supported by multiple S2G strategies. We further applied cS2G to provide a quantitative assessment of the “omnigenic hypothesis”^44–46^, concluding that the top 200 (1%) of ranked genes explained roughly half of the heritability linked to all genes; this implies that gene-level architectures in *cis* are largely driven by a relatively modest number of top genes.

Our framework, using polygenic analyses of disease heritability, is a substantial advance over previous approaches for evaluating S2G strategies using curated lists of disease-associated SNP-gene pairs^42,51^. In particular, curated lists of disease-associated SNP-gene pairs may contain SNPs whose causality has not been quantified (e.g. the Open Targets curated list^42^ uses sentinel SNPs) and may contain ascertainment biases. Experimentally validated enhancer-gene pairs^34,76,77^ provide an *in vitro* validation in a specific cell type for specific types of S2G strategies (i.e. enhancer-gene links), but this validation may not extend to *in vivo* disease contexts, which may involve cell types and cell states that are different from those assayed in validation experiments^27,28,34,76,77^; in particular, our genome-wide GWAS-based estimates of precision and recall for linking disease risk variants to disease genes are not comparable to the CRISPR-based estimates of precision and recall in K562 cells in Fig. 3 of ref. ^34^. Furthermore, unlike previous approaches, our framework provides a route to optimally combining S2G strategies, greatly improving precision and recall relative to individual strategies (Figure 2) as well as previously proposed combined strategies (Supplementary Table 5).

Our findings have several implications for downstream analyses. First, we recommend that GWAS fine-mapping studies employ cS2G to powerfully link fine-mapped SNPs to their target genes; we note that, as with previous S2G approaches, cS2G can be combined with similarity-based approaches leveraging genome-wide patterns of associated genes (as proposed in ref. ^51^). Second, our framework can be can be used to optimize (and combine) S2G strategies that may be developed in the future, including S2G strategies leveraging joint generation of single-cell ATAC-seq and single-cell RNA-seq data sets across a large number of tissues and cell-types; the development of new S2G strategies remains a key priority, as our cS2G strategy—despite its large improvement over other S2G strategies—attained a modest recall of 33%, implying that only 1/3 of causal disease SNPs can be functionally linked to their correct target genes. Third, our results highlight the advantages of enhancer-gene linking strategies such as EpiMap and ABC in future efforts to improve S2G strategies; their advantages include cost effective experiments targeting multiple cell-types (EpiMap and ABC provide links for 833 and 167 cell-types, respectively), and high potential for linking rare variants to genes. Fourth, our findings support the hypothesis that rare variant association studies^78,79^ will provide biological insights complementary to those of GWAS—both because we observed little overlap between genes underlying common variant and low-frequency variant gene-level architectures, and because we determined that low-frequency variant gene-level architectures were less polygenic (median ratio of 2.8); we expect these differences to be even more pronounced for rare variant architectures. Finally, cS2G can improve identification of gene sets that are enriched for disease heritability (although it has not been optimized for this specific purpose). For example, in contrast to the prevailing approach of applying S-LDSC^11^ to gene sets using ±100kb windows to define SNP annotations^50,80^, using cS2G to define SNP annotations produced larger heritability enrichments and standardized effect sizes (Supplementary Figure 10; also see ref. ^74^); we further note the importance of including appropriate SNP annotations in the model used by S-LDSC in analyses of enriched gene sets, in order to avoid biased enrichment estimates (see Methods and Supplementary Figure 11). Investigating the relative performance of different combined S2G strategies in analyses of gene sets that are enriched for disease heritability is a direction for future research; although we have focused here on S2G strategies defined using all available tissues/cell-types (see Supplementary Figure 3), tissue-specific S2G strategies may be preferred when analyzing gene sets reflecting genes that are specifically expressed in a particular tissue/cell-type^74^ (or when larger data sets become available; see below).

We note several limitations of our work. First, our definition of precision assumes that the Exon strategy has a precision of 1, but this is an approximation as the link between exonic SNPs and target genes is likely to be imprecise in some cases. Second, our estimates of precision had large standard errors for S2G strategies linking a limited fraction of SNPs to genes (Supplementary Figure 1 and Supplementary Table 5), such that evaluation of these S2G strategies was imprecise; however, for the cS2G strategy, estimates of precision (0.75, s.e. 0.06) and recall (0.33, s.e. 0.03) were reasonably precise. Third, we restricted each S2G strategy to the gene(s) with the highest linking score, as we observed that this led to slightly higher precision (Supplementary Figure 12). This does not reflect biological reality, in which a regulatory element may target more than one gene^18,37,38^; refinements to this choice are a direction for future research. Fourth, we included all available tissues and cell types for constituent S2G strategies of cS2G, as we observed that this led to higher precision (Supplementary Figure 3), perhaps due to limited biosample size. However, as larger data sets become available, it may become practical to define disease-specific combined S2G strategies that restrict to disease-critical tissues and cell types, furthering the goal of pinpointing the causal cell-types of action of SNP-gene-disease triplets; these can be evaluated under our framework by meta-analyzing results of disease-specific combined S2G strategies across diseases/traits to obtain precise estimates of the precision and recall of a specific approach (Supplementary Figure 3). Finally, our analyses using cS2G (and its constituent S2G strategies^11,18,19,26,30,33,34,36–38,47^) pertain exclusively to SNP-gene pairs *in cis*, and do not capture *trans* effects, which may contribute substantially to disease omnigenicity^46^; in particular, this may explain why the disease heritability linked to genes in *cis* using the cS2G strategy (*h*^2^_*gene*_) represents only roughly half of total SNP-heritability (*h*^2^). Despite these limitations, our results convincingly demonstrate both the advantages of using our polygenic framework to evaluate and combine S2G strategies, and the effectiveness of using our cS2G strategy to pinpoint disease genes.

## Data Availability

The List of 19,995 genes, summary statistics of the 63 independent traits, training and validation critical gene sets, S2G and cS2G strategies, SNP annotations, predicted causal SNP-disease pairs from UK Biobank fine-mapping analyses and from the NHGRI-EBI GWAS catalog, and heritability causally explained by SNPs linked to each gene have been made publicly available at https://alkesgroup.broadinstitute.org/cS2G.

## Acknowledgments

We thank X. Jiang, C. Boix and M. Kellis for helpful discussion. S.G. is funded by NIH grant R00 HG010160. A.L.P. is funded by NIH grants U01 HG009379, R01 MH101244, R37 MH107649, R01 MH115676 and R01 MH109978.

## Code Availability

Code will be made publicly available at https://alkesgroup.broadinstitute.org/cS2G prior to publication.

## Data Availability

GTeX *cis*-eQTLs https://storage.googleapis.com/gtex_analysis_v8/single_tissue_qtl_data/GTEx_Analysis_v8_eQTL.tar eQTLGen blood *cis*-eQTLs https://molgenis26.gcc.rug.nl/downloads/eqtlgen/cis-eqtl/2019-12-11-cis-eQTLsFDR-ProbeLevel-CohortInfoRemoved-BonferroniAdded.txt.gz

Roadmap enhancer-gene linking http://www.biolchem.ucla.edu/labs/ernst/roadmaplinking/RoadmapLinks.zip EpiMap enhancer-gene linking https://personal.broadinstitute.org/cboix/epimap/links/links_corr_only/

PCHi-C Jung links https://www.ncbi.nlm.nih.gov/geo/download/?acc=GSE86189&format=file&file=GSE86189%5Fall%5Finteraction%2Epo%2Etxt%2Egz and https://www.ncbi.nlm.nih.gov/geo/download/?acc=GSE86189&format=file&file=GSE86189%5Fall%5Finteraction%2Epp%2Etxt%2Egz

PCHi-C Javierre blood links https://ars.els-cdn.com/content/image/1-s2.0-S0092867416313228-mmc4.zip

Open Targets links ftp://ftp.ebi.ac.uk/pub/databases/opentargets/genetics/190505/variant-index/

GWAS catalog https://www.ebi.ac.uk/gwas/api/search/downloads/full

Open Targets SNP-gene pairs https://raw.githubusercontent.com/opentargets/genetics-gold-standards/master/gold_standards/processed/gwas_gold_standards.191108.tsv

SNP-gene pairs from ref.^51^ https://urldefense.proofpoint.com/v2/url?u=https-3A_www.dropbox.com_s_kz2c49rpm2yanf5_all-5FbyCS-5Frev1.txt-3Fdl-3D0&d=DwMFaQ&c=WO-RGvefibhHBZq3fL85hQ&r=pj2hZETq-6Xv2-wuSquXm871XqnKfXGPV5duZ9gf88w&m=IWrDyJkE3HPLhLS3pLXW8e80amAyxaNTtTw2ULHrbLA&s=A8lWwTxhGFCV0avT0-G3s0X7Cs1TXlFqvPx8woQ6iiU&e=

## Methods

### SNP-to-gene (S2G) strategies

A SNP-to-gene (S2G) linking strategy *k* is defined as an assignment of a linking score *ψ*_*k,j,g*_ between each SNP *j* and zero or more candidate target genes *g*, such that each SNP has a sum of linking scores ≤1 (we allowed Σ_*g*_ *ψ*_*k,j,g*_ < 1 to allow for incomplete SNP-to-gene linking; see below). We considered only links related to a list of 19,995 genes, including 17,871 protein-coding genes, that pass our quality control procedure (see Data Availability). Specifically, we selected genes in Ensembl^49^, GENCODE^47^, and RefSeq^48^ databases that have a unique identifier in all the datasets, overlapping starting and ending gene positions, and similar strand information. We verified that restricting our analysis to 17,871 protein-coding genes (instead of 19,995 protein-coding and non-protein-coding genes) had little impact on our results (Supplementary Figure 13).

We considered 50 S2G strategies (Table 1 and Supplementary Table 2); in each case we first considered raw linking values *A*_*k,j,g*_, which we next converted into linking scores *ψ*_*k,j,g*_ (see below):

#### Exon

for each gene, the list of each exons was extracted from the GENCODE database^47^. We added 20bp flanking windows between the exons to include splice regions.

#### Promoter

for each gene, we selected the list of TSSs of different transcripts from the Ensembl dataset, added a +/-1kb regions around them, intersected with promoter annotations from the baseline model^11^ (including promoters from refs.^29,81,82^) and from Roadmap^30^, and removed regions overlapping exons or splice regions.

#### Gene body

for each gene, we selected the minimum starting position and maximum ending position across Ensembl, GENCODE, and RefSeq databases.

#### Gene±100kb

for each gene, we added a +/-100kb window around its gene body.

Closest TSS and Closest *i*^*th*^ TSS (20 strategies): for each SNP, we defined its closest, second closest, and up to 20^th^ closest TSS based on physical distance with the TSSs of different transcripts from the Ensembl dataset.

#### Distance constrained closest TSS (7 strategies)

we only linked SNPs for which closest TSS was less than 1kb away, between 1kb and 5kb away, between 5kb and 10kb away, between 10kb and 50kb away, between 50kb and 100kb away, between 100kb and 500kb away, and between 500kb and 1,000kb away.

#### GTEx *cis*-eQTL

we used GTEx v8 significant variant-gene associations for each of the 54 cell-types (17,382 samples in total), kept the minimum variant-gene association *P* value when a variant was link to a gene in multiple cell-types, and used -log10 of this *P* value as raw linking value.

#### GTEx blood/immune *cis*-eQTL

same as GTEx *cis*-eQTL, but by restricting eQTLs from three blood/immune cell-types (i.e. whole blood, spleen and EBV-transformed lymphocytes).

#### GTEx fine-mapped *cis*-eQTL

we fine-mapped the GTEx v8 *cis*-eQTLs of each gene in each tissue as in ref.^26^, selected the SNPs with a causal posterior probability (CPP) ≥0.05, kept the maximum variant-gene CPP when a variant was fine-mapped to a gene in multiple tissues, and used corresponding CPP as raw linking value.

#### GTEx blood/immune fine-mapped *cis*-eQTL

same as GTEx fine-mapped *cis*-eQTL, but by restricting fine-mapped cis-eQTL from three blood/immune cell-types.

#### eQTLGen blood *cis*-eQTLs

we used eQTLGen statistically significant *cis*-eQTLs in blood (31,684 individuals), and used corresponding -log10 *P* value as raw linking value.

#### eQTLGen fine-mapped blood *cis*-eQTL

we fine-mapped the eQTLGen *cis*-eQTLs of each gene in each tissue as in ref.^26^, selected the SNPs with a CPP ≥0.05, and used corresponding CPP as raw linking value.

#### Roadmap enhancer-gene linking

we used linked Roadmap enhancers^29,30,32^ based on expression-enhancer activity correlation across 127 cell-types, kept the maximum correlation when an enhancer was linked to a gene in multiple tissues, and used corresponding correlation as raw linking value.

#### Roadmap blood/immune enhancer-gene linking

same as Roadmap enhancer-gene linking, but by restricting enhancers from 27 blood/immune cell-types.

#### EpiMap enhancer-gene linking

we used linked EpiMap enhancers^29,37^ based on expression-enhancer activity correlation across 833 cell-types, kept the maximum correlation when an enhancer was linked to a gene in multiple tissues, and used this correlation as raw linking value.

#### EpiMap blood/immune enhancer-gene linking

same as EpiMap enhancer-gene linking, but by restricting enhancers from 85 blood/immune cell-types.

#### Activity-by-Contact (ABC)

we used ABC links from ref.^38^, kept the maximum ABC score across the 167 cell-types when an enhancer was interacting with a gene in multiple cell-types, and used this score as raw linking value.

#### ABC blood/immune

same as ABC, but by restricting elements from 69 cell-types with blood/immune tissue/cells.

#### Closest TSS (Hi-C)

for each SNP, we defined TSS with the highest Hi-C intensity (averaged across 10 cell-types^34^) with the TSSs of each gene as in ref.^34^. We labeled this strategy closest TSS (Hi-C) as it gave similar results to the closest TSS strategy based on physical distance (Supplementary Table 5).

#### Hi-C distance

we linked each SNP to its surrounding genes using Hi-C intensity (averaged across 10 cell-types^34^) with the TSS of each gene, as in ref.^34^.

#### Jung PCHi-C

we used PCHi-C links from ref.^35^, kept the minimum interaction *P* value across the 27 cell-types when a genomic region was interacting with a gene in multiple cell-types, and used corresponding -log10 *P* value as raw linking value.

#### Javierre PCHi-C blood

we used PCHi-C links from ref.^31^, kept the maximum CHiCAGO score across the 17 cell-types when a genomic region was interacting with a gene in multiple cell-types, and used corresponding CHiCAGO score as raw linking value.

#### Cicero blood/basal

we used the enhancer-promoter links from ref. (data obtained through personal request to the authors), and used the enhancer-promoter correlation as raw linking value.

#### GeneHancer

we used the GeneHancer dataset^39^ and used their enhancer scores as raw linking values.

#### Open Targets

we downloaded Open targets annotations, and weight each annotation as suggested in Open Targets Genetic website (i.e. 1.00 for variant effect predictor (VEP), 0.66 for expression and protein QTLs, and 0.33 for distance to TSS, PCHi-C, and DHS-promoter and enhancer-TSS interactions); we used these values as raw linking values.

For all but one strategy (Hi-C distance, see below), we converted raw linking values *A*_*k,j,g*_ (as defined above) into linking scores *ψ*_*k,j,g*_ such that each SNP has a sum of linking scores over genes being 0 or 1 (we note that linking score *ψ*_*k,j,g*_ should not be interpreted as probabilities). When an S2G strategy linked a SNP to multiple genes, we considered three different ways to convert raw linking values *A*_*k,j,g*_ (as defined above) into linking scores *ψ*_*k,j,g*_: for a given SNP *j* we 1) assigned to each gene *g* with *A*_*k,j,g*_ > 0 the same linking score, 2) assigned to each gene a linking score proportional to its raw linking value, and 3) kept the gene(s) with the highest linking score (if the linking value was similar for *G* genes, we assigned a score of 1/*G* to each of these genes). We compared the precisions obtained using these 3 approached by using our training critical gene set (Supplementary Figure 12). For Hi-C distance, we created *ψ*_*k,j,g*_ linking scores proportional to Hi-C intensities (i.e. *ψ*_*k,j,g*_ = *A*_*k,j,g*_/ Σ_*g*_ *A*_*k,j,g*_), unless the sum of the intensities across genes was below 0.2 (in that case *ψ*_*k,j,g*_ = *A*_*k,j,g*_/0.2), in order to have Σ_*g*_ *ψ*_*k,j,g*_ < 1 for SNPs *j* in gene deserts.

Correlations between the S2G strategies were computed on all SNP-gene links observed by at least one of 34 S2G strategies (we omitted 6^th^ closest TSS to 20^th^ closest strategy and Hi-C due to computational constraints) (Supplementary Table 1). We defined a subset of 13 independent S2G strategies (different from the 13 main strategies) with *r*^*2*^ < 0.1 when comparing *h*^*2*^ coverage, precision or recall estimates.

We note that the functionally informed S2G strategies are derived from functional assays with widely varying biosample sizes. For example, PCHi-C datasets used 17/27 cell-types, enhancer maps such as EpiMap or ABC used multiple functional assays for 127 and 833 cell-types, and cis-eQTLs such as GTeX and eQTLGen used 17,382 and 31,684 RNA-seq samples. Thus, our evaluation of S2G strategies should not be viewed as an evaluation of the underlying functional assays.

### Evaluation of S2G strategies

To evaluate each S2G strategy’s informativeness for pinpointing disease genes, we aimed to define and estimate parameters that correspond to an S2G strategy’s heritability coverage (proportion of total disease SNP-heritability (*h*^*2*^) that is linked to genes; *h*^*2*^ coverage), precision (proportion of linked disease *h*^*2*^ that is linked to the correct target gene), and recall (proportion of total disease *h*^*2*^ that is linked to the correct target gene).

First, we defined *h*^*2*^ *coverage* as the proportion of *h*^*2*^ explained by all SNPs linked to one or more genes (weighted by their linking scores):

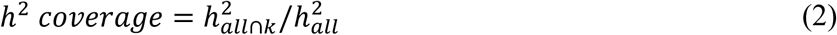

where 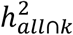 is the heritability explained by common SNPs linked all genes using *k*, and 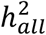is the heritability explained by common SNPs.

Second, we defined *precision* as the relative excess *h*^*2*^ enrichment of SNPs linked to a critical gene set (see below) vs. SNPs linked to all genes, as compared to the (gold-standard) Exon S2G strategy. More precisely, the precision of an S2G strategy *k* was defined as

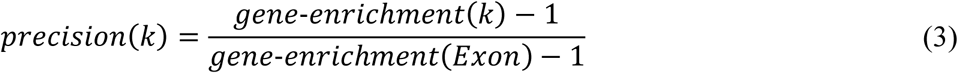

With

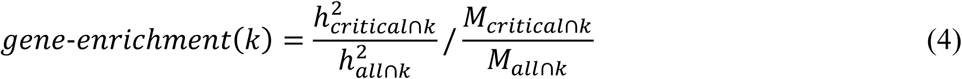

where 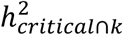 is the heritability explained by common SNPs linked to the critical gene set using *k, M*_*critical*∩*k*_ is the number of common SNPs linked to the critical gene set using *k*, and *M*_*all*∩*k*_ is the number of common SNPs linked to all genes using *k*. We note that this definition relies on the hypothesis that genes in the critical gene set are enriched for causal disease genes (as observed empirically, see below), and the hypothesis that the Exon S2G strategy is a perfectly precise strategy (even though it suffers from low *h*^*2*^ coverage). Third, we defined *recall* as the product of the *h*^*2*^ coverage and precision.

We estimated these quantities using polygenic analyses of disease heritability by applying stratified LD score regression (S-LDSC) with the baseline-LD model (v2.2)^11,52,53^ to 63 independent diseases and complex traits (average *N* = 314K; Supplementary Table 3), meta-analyzing results across traits. We analyzed SNP annotations for ∼10M SNPs with a minor allele count ≥5 in a 1000 Genomes Project European reference panel^54^. We jointly considered the 97 SNP annotations of the baseline-LD model v2.2 (refs.^52,53^), 50 S2G-derived SNP annotations constructed by restricting SNPs linked to genes of the critical gene set, and 30 S2G-derived SNP annotations constructed by restricting SNPs linked to all 19,995 genes (we did not include SNP annotations constructed using all genes for the 20 closest TSS S2G strategies, as these would include all SNPs), for a total of 177 SNP annotations. Jointly considering all these S2G-derived SNP annotations was crucial to maximize the accuracy of 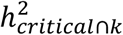 (Supplementary Figure 11). This strongly demonstrates the importance of including appropriate SNP annotations in the model used by S-LDSC in analyses estimating the proportion of heritability explained by enriched gene sets^74^, in order to avoid biased enrichment estimates. We note that precisions from preliminary analyses of Supplementary Figure 12 were estimated using the baseline-LD model and S2G-derived SNP annotations constructed from the Exon, Promoter, Gene body, Gene body +/- 100kb, and Closest TSS S2G strategies; using this restricted set of S2G strategies attenuated the bias of 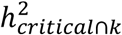.

We estimated values of *h*^*2*^ coverage and gene-enrichment for each disease/trait, estimated their standard errors using a genomic block-jackknife with 200 blocks, meta-analyzed results across the 63 independent traits using a fixed-effect meta-analysis, and used these values to estimate precision and recall. Precision and recall were estimated from meta-analyzed *h*^*2*^ coverage and gene-enrichment values (instead of meta-analyzing precision and recall across traits) to guarantee robust estimates. Precision and recall standard errors were estimated by using the 200 *h*^*2*^ coverage and gene-enrichment estimates from the block-jackknife procedure, but meta-analyzed using the same *h*^*2*^ coverage and gene-enrichment standard errors.

### Training and validation critical gene sets

Our definitions of precision and recall rely on a critical gene set. We used a non-trait-specific *training* critical gene set to construct an optimal combined S2G strategy, and trait-specific *validation* critical gene sets to evaluate the optimal combined S2G strategy while avoiding overfitting (for comparison purposes, we also used the validation critical gene sets to evaluate individual S2G strategies). Training and validation critical gene sets rely on information from exons and promoters to guarantee high-confidence SNP-gene links.

We defined a non-trait-specific training critical gene set as the top 10% of genes with the most highly constrained exons and conserved promoters. Specifically, for each gene we multiply its pLI score^83^ (estimating gene probability to be intolerant to loss-of-function mutations) by the fraction of bases of its promoter (defined using the Promoter S2G strategy) that is conserved (defined using 4 baseline-LD conserved SNP annotations^84,85^) (note that 17,554 out of our 19,995 genes had a pLI score). As we observed a correlation between this score and gene body length (*r* = 0.18), we created 10 bins of genes based on their gene body length, and selected the genes with the top 10% of this score (1,760 genes in total).

We defined the validation critical gene set for a given trait as the top 10% of genes ranked by the PoPS method^51^ (we note that PoPS gene scores are based on a leave-one-chromosome-out approach, implying that gene scores should be independent of their surrounding SNPs). By default, the initial step of PoPS is to apply MAGMA^86^ to compute gene-level association statistics, which relies on linking SNPs to genes using a gene body S2G strategy. To limit the impact of the gene body S2G strategy in our analyses, we modified PoPS to only link SNPs that are in exons or promoters (note that this led to nearly similar heritability enrichment and gene-enrichment values; Supplementary Table 4). 16,728 out of our 19,995 genes had a PoPS score, leading to validation critical gene sets with 1,673 genes.

The excess overlap between the training and validation critical gene sets was limited (median (across traits) overlap of 20% of genes in each gene set, vs. 10% expected by chance). We also observed substantial excess overlap of housekeeping genes (1,997 genes^87^) in the training critical gene set (357 genes; excess overlap = 2.0) and validation critical gene sets (median of 281 genes across 63 traits; median excess overlap = 1.7).

### Combining S2G strategies

We constructed combined S2G strategies as linear combinations of linking scores from *K* S2G strategies. Specifically, for each SNP *j* and gene *g* we computed a combined S2G linking score

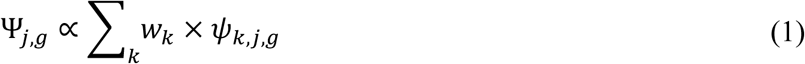

where *ψ*_*k,j,g*_ is the linking score between SNP *j* and gene *g* for S2G strategy *k*, and *w*_*k*_ is the weight associated to strategy *k*. We used Ψ_*j,g*_ = Σ_*k*_ *w*_*k*_ × *ψ*_*k,j,g*_ when Σ_*g*_ Σ_*k*_ *w*_*k*_ × *ψ*_*k,j,g*_ < 1, and Ψ_*j,g*_ = Σ_*k*_ *w*_*k*_ × *ψ*_*k,j,g*_ / Σ_*g*_ Σ_*k*_ *w*_*k*_ × *ψ*_*k,j,g*_ otherwise; allowing Σ_*g*_ Ψ_*j,g*_ to be < 1 allows to give small combined S2G linking scores to SNPs with linking score available only for imprecise S2G strategies. Here, we allowed weights *w*_*k*_ to have a maximum value of 100, to prioritize S2G strategies with higher precision in the case where two S2G strategies link the same SNP to different genes. For example, if we have an S2G strategy 1 with high precision and low recall and an S2G strategy 2 with reasonable precision and high recall, then assigning weights of 100 and 1 allows to create a combined S2G strategy that will leverage the high precision of S2G strategy 1 when a SNP is linked to different genes using S2G strategies 1 and 2, while maximizing recall using S2G strategy 2.

To estimate the optimal weights *w*_*k*_, we developed an optimization framework to identify the weights maximizing the recall while constraining precision (defined using the training critical gene set) to be ≥0.75. First, we computed for each SNP its expected per-SNP heritability by meta-analyzing across the 63 independent traits the S-LDSC regression coefficients estimated with the baseline-LD model and the 80 S2G-derived SNP annotations of the training gene set and all genes. Second, we defined a function taking as input a vector of weights *w*, computed for each SNP the expected per-SNP heritability linked to the critical gene set and to all genes using the 80 S2G-derived SNP annotations, and outputting precision and recall. Finally, we found the vector *w* maximizing recall while constraining precision to be ≥0.75. Specifically, we considered a grid of values for *w*, going from 0 to 2.5 with a 0.1 step, and the values 5, 7.5, 10, 25, 50, 75, and 100 (33 total values). We created a custom optimization framework that (a) starts by giving a weight of 0.1 to all S2G strategies investigated, (b) computes precision and recall by increasing a single value of *w* at a time (the higher weight on the grid), and keeps for next step the weight vector maximizing recall (constraining precision to be ≥0.75), (c) computes precision and recall by decreasing a single value of *w* at a time (the lower weight on the grid), and keeps for next step the weight vector maximizing recall (constraining precision to be ≥0.75), (d) computes precision and recall by randomly modifying a single value of *w* at a time, and keeps for next step the weight vector maximizing recall (constraining precision to be ≥0.75), and (e) restarts from (b) till the recall does not improve. We repeated this algorithm 5 times, and kept the weight vector providing the maximum recall. We note that we investigated 5 different optimization algorithms from the R software (methods Nelder-Mead, BFGS, CG, L-BFGS-B and SANN from the *optim* function), but none of them reached higher recall than our algorithm. When giving as a starting point the outputs of our custom algorithm, these 5 algorithms did not converge to significantly different weight vectors and recalls.

We note that we allowed weights to have a maximum value of 100, to prioritize S2G strategies with higher precision in the case where two S2G strategies link the same SNP to different genes. For example, in the case of our cS2G strategy (Supplementary Table 6), if a SNP is linked to gene A through the Exon S2G strategy (weight = 100), and to gene B through the Cicero S2G strategy (weight = 1), then the cS2G linking score is 100/101 for gene A (stronger evidence from Exon), and 1/101 for gene B. We note that weights of 10 and 0.1 for Exon and Cicero (rather than 100 and 1), would have assigned the same linking scores in the case of the SNP described above, but would have assigned lower linking scores to SNPs that are linked to genes only through Cicero. However, we note that fixing the weights of the 7 constituent S2G strategies of the cS2G strategy to the same value only slightly underperformed cS2G in both precision and recall (0.71 vs. 0.75 and 0.31 vs. 0.33, respectively; Supplementary Table 8), as expected given the low overlap of SNPs annotated by these 7 strategies (Supplementary Table 7).

### Curated SNP-gene pairs for validation

We verified that our estimates of precision and recall (based on polygenic analyses of disease heritability) were similar to independent definitions (not relying on critical gene sets or polygenic analyses) based on two curated SNP-gene pairs. First, we considered the list of linked sentinel SNP-gene pairs with the underlying genes validated with high confidence by Open Targets^42^. We selected SNP-gene-disease triplets defined with “high” confidence, selected sentinel SNPs with a minor allele count ≥5 in a 1000 Genomes Project European reference panel^54^, selected genes in our list of 19,995 genes, and kept unique SNP-gene pairs, leading to a total of 577 pairs. Second, we used linked fine-mapped SNP-gene pairs validated using nearby fine-mapped protein-coding variants^51^. After a similar control quality procedure, we retained 1,668 pairs. We estimated precision and recall by analyzing only the SNPs of the SNP-gene pairs, and by assuming that all SNP-gene pairs that are not on the list are false positives.

We note that the Open Targets dataset reports sentinel SNPs (rather than causal fine-mapped SNPs), which may be linked to different genes than causal SNPs, as we verified in analyses of UK Biobank traits (Supplementary Table 17). We also note that these two curated SNP-gene pairs were preferentially ascertained for disease-associated SNPs in which the target gene was the closest gene: indeed, we observed an unusually high proportion of SNP-gene pairs involving closest TSS genes with a small distance (< 10kb) to the closest TSS (57% and 67% for the two curated lists, vs. *h*^*2*^ coverage = 34% for the Closest TSS <10kb S2G strategy). Thus, we caution that curated disease-associated lists of linked SNP-gene pairs may be non-randomly ascertained.

### Leveraging the combined S2G strategy to pinpoint disease genes

We analyzed 9,670 predicted causal SNP-disease pairs with posterior inclusion probability (PIP) >0.50 from functionally informed fine-mapping of 49 UK Biobank diseases/traits using PolyFun + SuSiE^7,43^. For this purpose, we created S2G and cS2G strategies for 19M imputed UK Biobank SNPs with MAF ≥ 0.1% (we note that these analyses include both SNPs and indels, but we use the term SNP throughout the manuscript for simplicity). We predicted causal SNP-gene-disease triplets by restricting to SNPs that had a linked gene with cS2G linking score >0.5 (98% of the cS2G linked SNPs).

We further assigned a confidence score to each SNP-gene-disease triplet with a cS2G linking score >0.5 by multiplying their corresponding PIP and precision. To account for the excess of exon and promoter SNPs, we assigned the precision of Exon (i.e. 1.00) if the link was validated through the Exon S2G strategy, the precision of Promoter (i.e. 0.80) if the link was validated through the Promoter S2G strategy, and an estimated precision for SNPs that are not in exons or promoters (*precision*_*Other*_) otherwise. We estimated *precision*_*Other*_ for an S2G strategy *k* using the following formula

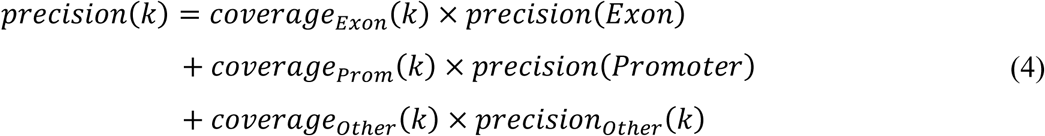

where *coverage*_*Exon*_, *coverage*_*Prom*_, and *coverage*_*Other*_ are the proportion of *h*^*2*^ coverage explained by SNPs in exons, promoters, and SNPs that are not in exons or promoters, respectively.

To predict the candidate cell-type of action for these triplets, we (i) focused on SNP-gene pairs provided by GTEx fine-mapped *cis*-eQTL, EpiMap enhancer-gene linking, and/or ABC, (ii) restricted to cell-types where the SNP-gene pair has been observed (out of 54, 833 and 167, respectively), and (iii) reported the cell-type with the most significant regression coefficient in an S-LDSC analysis conditioned to the baseline model (as performed in refs.^11,50^).

### Leveraging the combined S2G strategy to empirically assess disease omnigenicity

To estimate the heritability explained by SNPs linked to each gene *g* for 49 UK Biobank diseases/traits (per-gene heritability *h*_*gene,g*_^*2*^), we used PolyFun + SuSiE estimates of posterior mean squared causal effect sizes for 19M imputed SNPs with 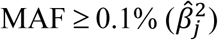 estimated on *N*=337K British UK Biobank samples^7,43^. First, we computed unadjusted per-gene heritability 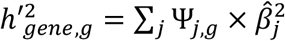, where Ψ_*j,g*_ is the cS2G linking score between SNP *j* and gene *g*. Then, we computed (adjusted) per-gene heritability 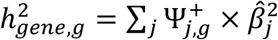, with 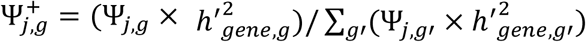 being a trait specific cS2G linking score. The motivation of this additional step is to improve the cS2G linking scores of SNPs linked to multiple genes by integrating evidence from 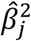 linked to a single gene; we note that this step changed per-gene heritability for only a small number of genes, as most SNPs linked using cS2G have large cS2G linking score (87% of the linked SNPs have a maximum cS2G linking score > 0.95) (see Supplementary Figure 14). We also estimated per-gene heritabilities linked to common SNPs (MAF≥5%) as 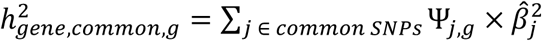, and per-gene heritabilities linked to low-frequency SNPs (0.1%≤MAF<5%) as 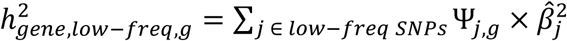.

To estimate the proportion of heritability linked to genes (*h*_*gene*_^2^) explained by genes with the top per-gene heritability, we partitioned genes into 8 gene sets ranked by per-gene heritability explained (top 100, 200, 500, 1,000, 2,000, 5,000, 10,000 and 19,995), and re-estimated the heritability linked to each ranked gene set by running S-LDSC with the baseline-LD model on summary statistics computed from *N*=122K European-ancestry UK Biobank samples that were distinct from the *N*=337K British UK Biobank samples used to estimate 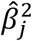 and 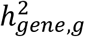 (in order to avoid winner’s curse, analogous to what was performed in ref.^7^). We defined *h*_*gene*_^2^ as the heritability linked to all 19,995 genes.

We estimated the effective number of causal genes (*G*_*e*_) for each trait using per-gene heritabilities and the formula of ref.^73^. Specifically, we defined *G*_*e*_ = 3*G*/*κ*, with G the total number of genes and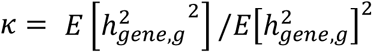. We extended this formula to per-gene heritabilities linked to common and low-frequency SNPs to estimate *G*_*e,common*_ and *G*_*e,low-frea*_, respectively. Per-gene heritabilities were directly estimated on the *N*=337K samples used for fine-mapping as we observed that the impact of winner’s curse on our analyses was minimal.

