## Supplementary Material for "Combining SNP-to-gene linking strategies to pinpoint disease genes and assess disease omnigenicity"

### Supplementary Table Legends

**Supplementary Table 1: Correlations between 34 SNP-to-gene (S2G) strategies.** We report the correlation between the linking scores across 34 S2G strategies (we omitted $6^{th}$ closest TSS to $20^{th}$ closest strategy and Hi-C due to computational constraints). Correlations were computed on all SNP-gene links observed by at least one of the 34 SG strategies (i.e. most of the observed scores were 0 for the two S2G considered in the correlation computation). We observed low concordance between the S2G strategies: the average correlation is 0.10 across the 34 S2G strategies, 0.09 across the 13 main strategies, and 0.05 across the 10 main functionally informed strategies.

**Supplementary Table 2: Description of the 50 SNP-to-gene (S2G) strategies.**

**Supplementary Table 3: List of 63 diseases/traits used to estimate $h^2$ coverage, precision and recall**. We defined a list of 63 summary statistics with independent association data (labeled as independent traits) by excluding genetically correlated traits in overlapping samples by measuring the intercept of cross-trait LD score regression[1], as previously described[2]; for traits with summary statistics computed from UK Biobank data, we also excluded traits with a squared genetic correlation[3] greater than 0.1 (similar to the squared phenotypic correlation threshold used in ref.[4]). The 63 datasets included six traits that were duplicated in two different datasets (genetic correlation of at least 0.9). Thus, we analyzed 57 independent diseases and complex traits. Traits were prioritized using the z-score for nonzero heritability computed using S- LDSC with the baseline-LD model (minimum of 6, as in ref.[5]). We also considered the 11 autoimmune diseases and blood cell traits, as in refs.[6,7].

**Supplementary Table 4: Estimates of $h^2$ enrichment and gene enrichment for the validation and training critical gene sets for 50 S2G strategies**. We report the $h^2$ enrichment, gene enrichment, and corresponding standard errors, meta-analyzed across 63 traits, for 50 S2G strategies, for both the (trait-specific) validation critical gene sets and the training critical gene set. $h^2$ enrichment is defined as the proportion of common variant heritability linked to the critical gene set, divided by the proportion of common SNPs linked to the critical gene set. Gene enrichment is defined as the fraction of common variant heritability linked to the critical gene set and to all genes, divided by the fraction of common SNPs linked to the critical gene set and to all genes. We also report estimates for validation critical gene sets constructed using default PoPS score (i.e. creating gene-level association statistics using gene body S2G strategy, rather than using Exon and Promoter S2G strategies); results were similar to the default validation critical gene sets.

**Supplementary Table 5: Estimates of $h^2$ coverage, precision and recall for the validation and training critical gene sets for 50 S2G strategies and the cS2G strategy**. We report the $h^2$ coverage, precision and recall, and corresponding standard errors, meta-analyzed across 63 traits, for 50 S2G strategies and the combined cS2G strategy, for both the (trait-specific) validation critical gene sets and the training critical gene set.

**Supplementary Table 6: Weights of constituent S2G strategies in the combined cS2G strategy**. We report the weights of each constituent S2G strategy in the combined cS2G strategy. We allowed weights to have a maximum value of 100, to prioritize S2G strategies with higher precision in the case where two S2G strategies link the same SNP to different genes. For example, if a SNP is linked to gene A through the Exon S2G strategy (weight = 100), and to gene B through the Cicero S2G strategy (weight = 1), then the cS2G linking score is 100/101 for gene A (stronger evidence from Exon), and 1/101 for gene B. We note that weights of 10 and 0.1 for Exon and Cicero (rather than 100 and 1), would have assigned the same linking scores in the case of the SNP described above, but would have assigned lower linking scores to SNPs that are linked to genes only through Cicero.

**Supplementary Table 7: Overlap of SNP-gene links between the constituent strategies of cS2G**. For each constituent strategy of cS2G (row), we report its fraction of links that are also validated by another S2G strategy (column). For example, 4.6% of links from the GTEx fine-mapped *cis*-eQTL strategy are also in the Exon strategy, and 17.6% of links from the Exon strategy are also in the GTEx fine-mapped *cis*-eQTL strategy. All the numbers are restricted to links involving common SNPs.

**Supplementary Table 8: Estimates of $h^2$ coverage, precision and recall for 4 different combined S2G strategies**. We report the $h^2$ coverage, precision and recall, and corresponding standard errors, meta-analyzed across 63 traits for 4 combined cS2G strategy. First, we considered our cS2G strategy. Second, we considered a combined S2G strategy with the same 7 S2G strategies than cS2G, but give them the exact same weight (i.e. 1) (cS2G – same weights). Third, we considered a combined S2G strategy where we linked all the SNPs linked by the cS2G (22% of common SNPs) to the gene with the closest TSS (cS2G – Closest TSS). Finally, we considered a combined S2G strategy maximizing recall (with precision > 0.75) when including all 50 S2G strategies (see Supplementary Table 9 for the 8 selected S2G strategies) (cS2G – 50 S2G). We report values estimated using both validation and training critical gene sets. For cS2G and cS2G – 50 S2G, we also report values estimated during the optimization algorithm using training critical gene sets. We note that the precision of the combined S2G strategies in the training critical gene set tend to be large (>0.85), which is very likely due to higher precision for some constituent S2G strategies in the training critical gene set compared to the validation critical gene set (such as promoter, Closest TSS (1kb-5kb), or GTEx fine-mapped *cis*-eQTL; see Supplementary Table 5).

**Supplementary Table 9: Combined S2G strategy obtained when including all 50 S2G strategies**. We report the selected S2G strategies and their corresponding weights that maximize recall (with precision > 0.75) when including all 50 S2G strategies. The resulting combined strategy included 8 S2G strategies: 4 that were included in our primary cS2G strategy (Exon, Promoter, eQTLGen blood fine-mapped *cis*-eQTL, GTEx fine-mapped *cis*-eQTL), as well as EpiMap and ABC restricted to blood and immune cell-types and tissues, Closest TSS (1-5kb), and GTEx all *cis*-eQTL.

**Supplementary Table 10: F1 scores of 50 S2G strategies and the combined strategy obtained when maximizing the F1 score**. We report the *F1 score*, the harmonic mean of precision and recall[8], for the 50 S2G strategies and a combined strategy maximizing the F1 score in the training critical gene set (cS2G-F1). cS2G-F1 contains 5 out of 7 S2G strategies of cS2G (all but EpiMap and ABC). F1 scores were computed based on precision and recall estimated on the validation critical gene sets, and meta-analyzed across 63 traits. Strategies are ranked based on their F1 score. We observed that cS2G-F1 optimizes the F1 score over the 50 SG strategies, showing that our framework is robust to the quantity to maximize. For comparison purposes, we also report the F1 score of the cS2G strategy and observed that cS2G F1 score is slightly higher than cS2G-F1 F1 score. This result may be due to precision and recall heterogeneity across the training and validation critical gene sets, as cS2G-F1 maximizes the F1 score over cS2G during the optimization procedure (0.49 vs. 0.47 for cS2G).

**Supplementary Table 11: Combined S2G strategies using different diseases/traits for training and validation**. We split the set of 63 diseases/traits in 2 ($1^{st}$ half and $2^{nd}$ half), built a combined S2G strategy using each of those (cS2G - 1st half, and cS2G - 2nd half, respectively), and report their $h^2$ coverage, precision and recall, and corresponding standard errors, meta-analyzed across each set of diseases/traits. cS2G - 1st half includes all the constituent S2G strategies of cS2G except EpiMap. cS2G - 2nd half includes all the constituent S2G strategies of cS2G except ABC and Cicero blood/basal. In all scenarios we observed that the combined strategies (cS2G - 1st half and cS2G - 2nd half) have a high precision (>0.76) and recall (>0.30). Note that in all scenarios, precision and recall were higher when using the training critical gene set than when using the validation critical gene set.

**Supplementary Table 12: Causal SNP-gene-disease triplets predicted by application of cS2G strategy to 9,670 fine-mapped SNP-disease pairs.** We report cS2G predictions for 9,670 predicted causal SNP-trait pairs with a posterior inclusion probability (PIP) > 0.50 from functionally informed fine-mapping of 49 UK Biobank diseases/traits[9,10]. Using cS2G linking scores >0.5, we predicted 7,111 causal SNP-gene-disease triplets (see column "In 7111") and report their corresponding confidence score. We also predicted 2,163 triplets involving distal regulatory fine-mapped SNPs that were not in the gene body (or promoter) of the target gene (see column "In 2163"), of which 532 were supported by at least 2 of the functionally informed constituent S2G strategies used by cS2G (see column "In 532"). We report the cS2G linking score as well as the score before normalization (column cS2G score*). We also report for each SNP-gene pair the annotations of the 7 constituent S2G strategies.

**Supplementary Table 13: cS2G strategy predicts more correct SNP-gene-disease triplets than other S2G strategies.** We report for the 13 main S2G strategies and the cS2G strategy the number of inferred SNP-gene-disease triplets from 9,670 predicted causal SNP-trait pairs with a posterior inclusion probability (PIP) > 0.50 from functionally informed fine-mapping of 49 UK Biobank diseases/traits[9,10]. For each strategy, we report the number of unique SNPs and genes in all the triplets, the mean PIP across all the triplets, the mean confidence score across all the triplets, and the number of correct SNP-gene-disease triplets (obtained by multiplying the number of inferred triplets by the mean confidence score). We observed that the cS2G links at least 1.6 times more unique genes and predict at least 2.0 times more correct SNP-gene-disease triplets than any of the other 10 functionally informed S2G strategies.

**Supplementary Table 14: Number of unique fine-mapped SNPs linked to each of the 3,401 unique genes in the 7,111 predicted SNP-gene-disease triplets.** In many instances, multiple causal SNPs were linked to the same gene. For example, 119 genes were each linked to at least 5 fine-mapped SNPs, illustrating that a single gene can be causal for different diseases/traits using different causal SNP-gene links.

**Supplementary Table 15: Cell-types identified by the functionally informed S2G strategies used by cS2G in the 4 examples of high-confidence SNP-gene-disease triplets identified by cS2G.** For the 6 SNPs involved in the 4 examples of Figure 5, we report all the cell-types identified by the functionally informed S2G strategies used by cS2G, their linking score, and their *P* value in LDSC-SEG analyses. Results are ordered by LDSC-SEG *P* value significance.

**Supplementary Table 16: cS2G predictions for a curated list of 577 sentinel SNP-gene pairs with genes curated with high confidence by Open Targets**. For each SNP in this list, we report the genes targeted by cS2G, their score, their corresponding annotations, and if the SNP-gene pair has been validated by Open Targets (column Validated). We also report the validated target gene if the cS2G lining score was 0. Note that the validated column has values different to 1 if multiple genes were assigned for one causal SNP. We warn here that while the causal gene has been validated with high confidence, the causal SNP(s) might be less confident as none of the 577 examples nominated causal SNPs using rigorous fine-mapping and/or functional follow-up, thus impacting the precision of the linking strategies. 356 of the 577 causal SNPs had a linked gene with a cS2G linking score >0.5, enabling us to predict the target gene. 205 of 356 predicted genes matched the Open Targets gene (precision = 205/356 = 0.58; we note that this precision definition, restricted to links with a cS2G linking score >0.5, is different from the one used in Supplementary Figure 5, which weights links by their cS2G linking score), and 205 of 577 causal SNP-gene-disease triplets were correctly identified (recall = 205/577 = 0.36) (see also Supplementary Figure 5). Of these 205 triplets, 178 involved the gene with closest TSS, and 88 (resp. 20) involved variants in exons (resp. promoters), illustrating that curated SNP-gene-disease triplets that can be validated using functionally informed S2G strategies are preferentially ascertained for triplets in which the target gene is the gene with the closest TSS and/or implicated by a high-confidence S2G strategy. Of the 97 SNP-gene-disease triplets involving distal regulatory variants (defined here as not lying in exons or promoters), only 31 were functionally supported by at least 2 of the 5 remaining functionally informed S2G strategies.

**Supplementary Table 17: Sentinel SNPs are linked to different genes than causal SNPs in analyses of fine-mapped UK Biobank traits.** We found 114 SNP-gene-trait triplets with the underlying genes validated with high confidence by Open Targets[11] and with available fine-mapping results in our analyses of 49 UK Biobank traits. For each triplet, we report the sentinel SNP, gene and trait defined by the Open Target list (4 first columns), the trait ID in our UK Biobank analyses (5th column), if the SNP-gene pair was validated by cS2G (6th column), and the posterior inclusion probability (PIP) in our fine-mapping analyses (7th column). 58/114 pairs were validated using cS2G (precision = 51%). The mean and median PIP of the 58 validated pairs were 0.260 and 0.045, respectively, against 0.055 and 0.004 for 56 unvalidated pairs (*P* Wilcoxon test = $1.6 \times 10^{-3}$). The precision is 93% (resp. 71%) when restricted to the 14 (resp. 41) triplets with PIP >0.50 (resp. >0.05). These results highlight that sentinel SNPs that are fine-mapped are more likely to be linked to the accurate target gene using cS2G, and potentially explain the discrepancy between the precision estimated using 577 Open Targets pairs (58%) and our estimated precision (75%).

**Supplementary Table 18: List of 49 UK Biobank diseases/traits used to empirically assess the omnigenic model**. We used the set of 49 traits and 16 independent traits as in ref. [10] (also same as in the fine-mapping analyses). We report the sample size used to estimate posterior mean squared causal effect sizes of genome-wide SNPs (*N*=337K British UK Biobank samples), and the sample size used to compute the summary statistics on European-ancestry UK Biobank samples that were distinct from the *N*=337K British UK Biobank samples (*N*=122K) for S-LDSC analyses.

**Supplementary Table 19: Numerical results of assessment of disease omnigenicity using cS2G**. We report the proportion of heritability ($h^2$) and the proportion of heritability linked to genes ($h^2_{gene}$) explained by the top 100, 200, 500, 1,000, 2,000, 5,000, 10,000 and all (19,995) genes using different linking strategies (cS2G and Closest TSS) (see Figure 5a). The top genes were defined using posterior mean squared causal effect sizes estimated on *N*=337K British UK Biobank samples, and reported proportions were estimated using S-LDSC on the N=122K samples and meta-analyzed across 16 independent traits. For the cS2G, we also report the proportion of heritability linked to genes ($h^2_{gene}$) estimated using posterior mean squared causal effect sizes estimated on the *N*=337K samples; we observed that from 200 genes, $h^2_{gene}$ S-LDSC estimates (based on *N*=122K samples) were not significantly different from the estimates directly based on the *N*=337K samples, implying minimal effects of winner's curse for a small number of genes.

**Supplementary Table 20: Heritability explained by top genes with the highest per-gene heritabilities for each disease/trait**. For each of the 49 traits, we report the proportion of heritability ($h^2$) and the proportion of heritability linked to genes ($h^2_{gene}$) explained by the top 100, 200, 500, 1,000, 2,000, 5,000, 10,000 and all (19,995) genes using the cS2G strategy.

**Supplementary Table 21: Estimates of the effective number of causal genes**. For each of the 49 traits, we report its effective number of causal SNPs ($M_e$) and causal genes ($G_e$) (see Figure 5b), its effective number of causal genes explained by common variants ($G_{e,common}$) and low-frequency variants ($G_{e,low\text{-}frequency}$) (see Figure 5c), its correlation between per-gene heritability explained by common and low-frequency variants across all the genes ($r_{20K}$), its correlation between per-gene heritability explained by common and low-frequency variants restricted to genes in the top 200 (i.e. 1%) of per-gene heritability explained by common and low-frequency variants ($r_{top200}$), and the shared number of genes in the top 200 (i.e. 1%) of per-gene heritability explained by common and low-frequency variants ($shared_{top200}$). We also report the median values across 16 independent traits.

**Supplementary Table 22: Top genes contributing to both common and low-frequency variant heritability linked to genes.** Across all 49 traits, we report the 19 triplets (13 unique genes) where the gene is in the top 3 genes contributing to the common and low-frequency variant heritability linked to genes ($h^2_{gene,common}$ and $h^2_{gene,low\text{-}freq}$, respectively). We note that our results include *CDKN1C* for type 2 diabetes, further validating *CDKN1C* as the causal gene at this locus.

#### Supplementary Figures

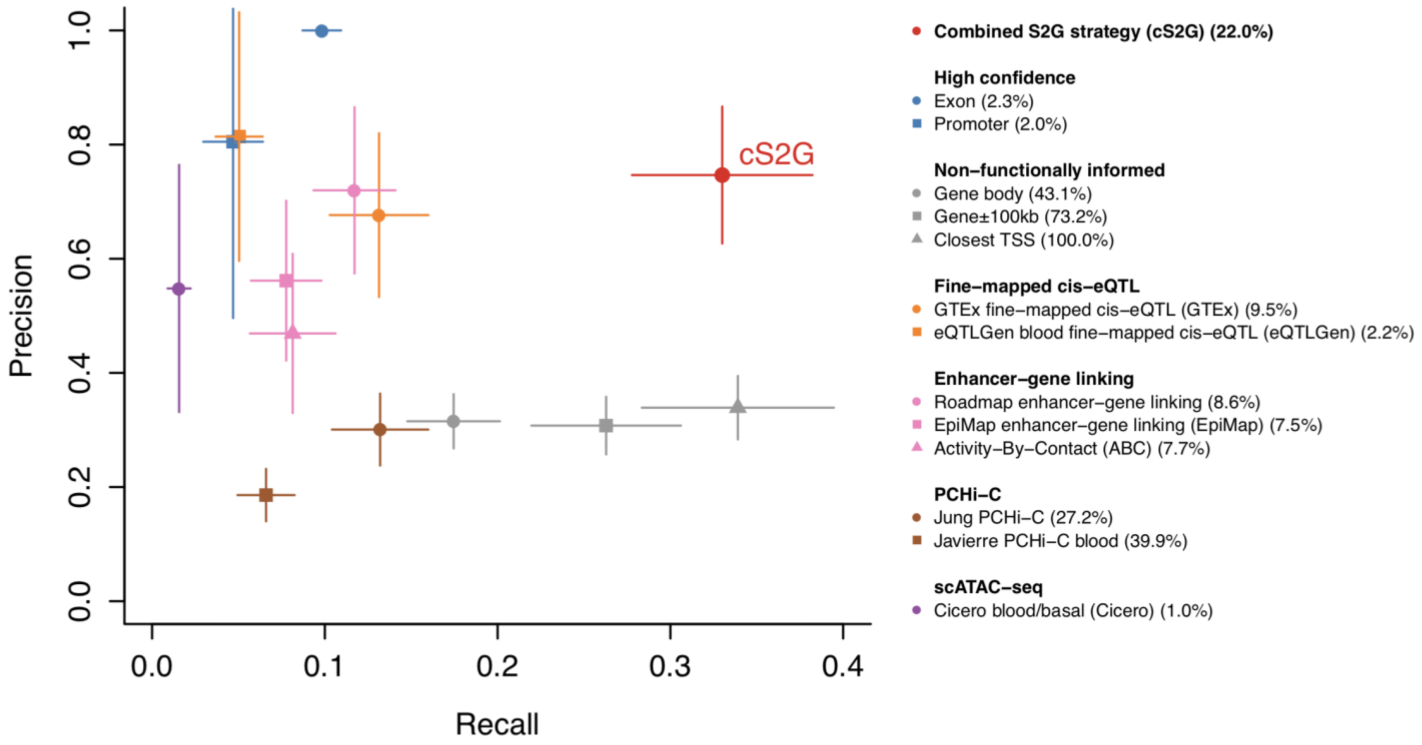


**Supplementary Figure 1: Accuracy (with standard error) of individual S2G strategies and combined S2G (cS2G) strategy.** Reported results are identical to Figure 2, except that they include estimated standard errors. Our estimates of precision have large standard errors for S2G strategies linking a limited fraction of SNPs to genes; however, for the cS2G strategy, estimates of precision (0.75, s.e. 0.06) and recall (0.33, s.e. 0.03) were reasonably precise.

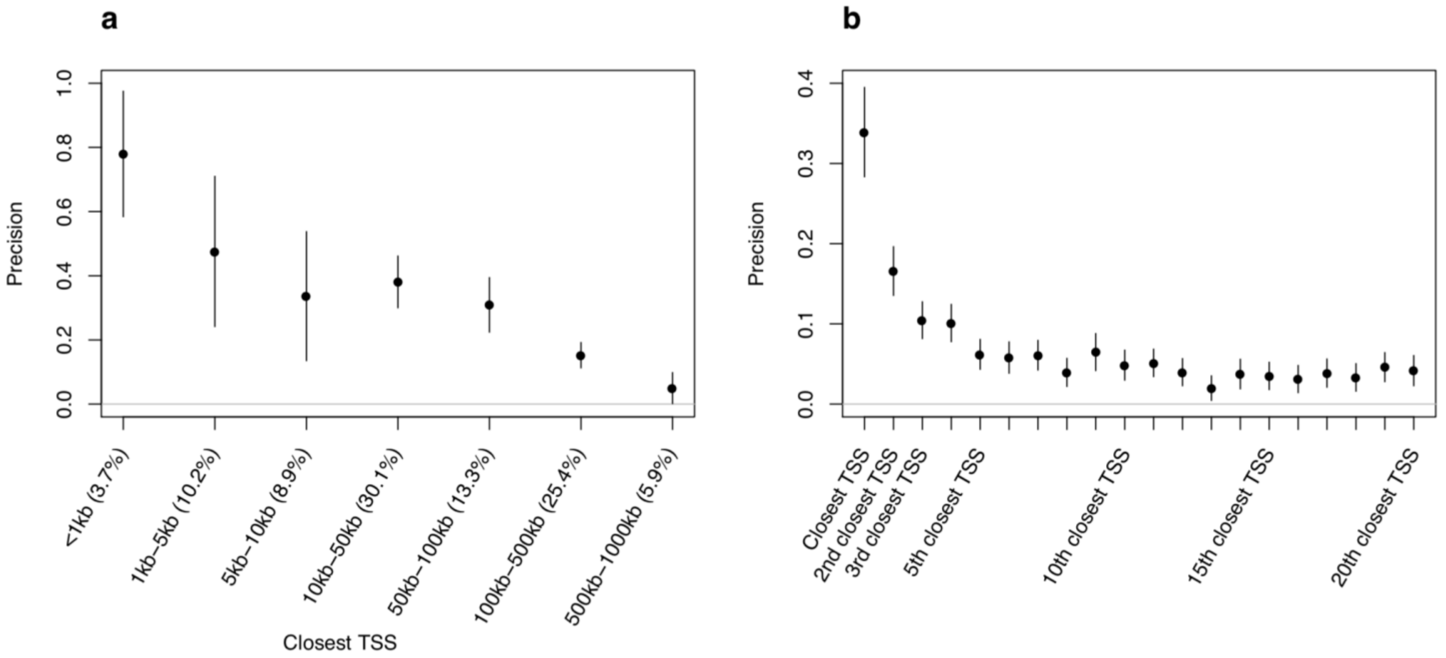


**Supplementary Figure 2: Precision of 27 S2G strategies based on physitical distance to TSS.** We report precision of the closest TSS strategy as a function of the distance between a SNP and its closest TSS **(a)** (numbers between parentheses represent the fraction of common SNPs linked by the strategy), and the precision of the $i^{th}$ closest TSS (each strategy links 100% of the SNPs) (**b**). The mean value of 0.043 for 6th-20th closest TSS suggests that genes located relatively close to causal disease genes have a slightly elevated probability of being causal. Numerical results including values of recall and corresponding standard errors are reported in Supplementary Table 5.

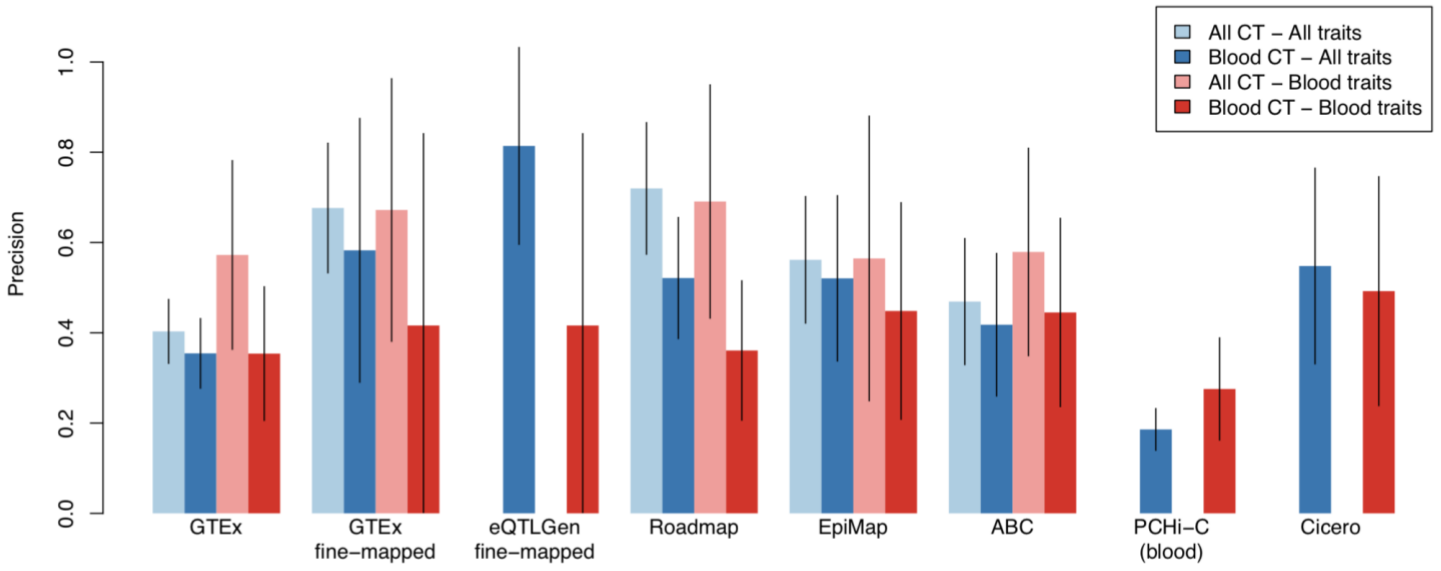


**Supplementary Figure 3: Precision of functional S2G strategies using all available cell-types and tissues or restricted to blood and immune cell-types and tissues.** We report the precisions and their 95% confidence intervals of functional S2G strategies built using either all available cell-types and tissues (All CT; in light color) and/or blood and immune cell-types and tissues (Blood CT; in dark color) estimated on 63 independent traits (All traits; in blue) and 11 blood cell traits and autoimmune diseases (Blood traits; in red) (UK Biobank all autoimmune diseases, Crohn's Disease, Rheumatoid Arthritis, Ulcerative Colitis, Lupus, Celiac, Platelet Count, Red Blood Cell Count, Red Blood Cell Distribution Width, Eosinophil Count, White Blood Cell Count; see Supplementary Table 3). We considered 5 S2G strategies with data available for cell-types and tissues: GTEx *cis*-eQTLs (GTEx), GTEx fine-mapped *cis*-eQTL (GTEx fine-mapped), Roadmap enhancer-gene linking (Roadmap), EpiMap enhancer-gene linking (EpiMap), and Activity-By-Contact (ABC). We considered 3 S2G strategies with data available only for blood and immune cell-types and tissues: eQTLGen fine-mapped blood *cis*-eQTL (eQTLGen fine-mapped), PCHi-C (blood), and Cicero blood/basal (Cicero). We observed 1) that S2G strategies using data from all cell-types and tissues were more precise than S2G strategies restricted to blood and immune cell-types and tissues in both analyses of all traits (light blue vs. dark blue) and blood cell traits and autoimmune diseases (light red vs. dark red), and 2) that S2G strategies using data from blood and immune cell-types and tissues are more precise in all traits than in blood cell traits and autoimmune diseases (dark blue vs. dark red).

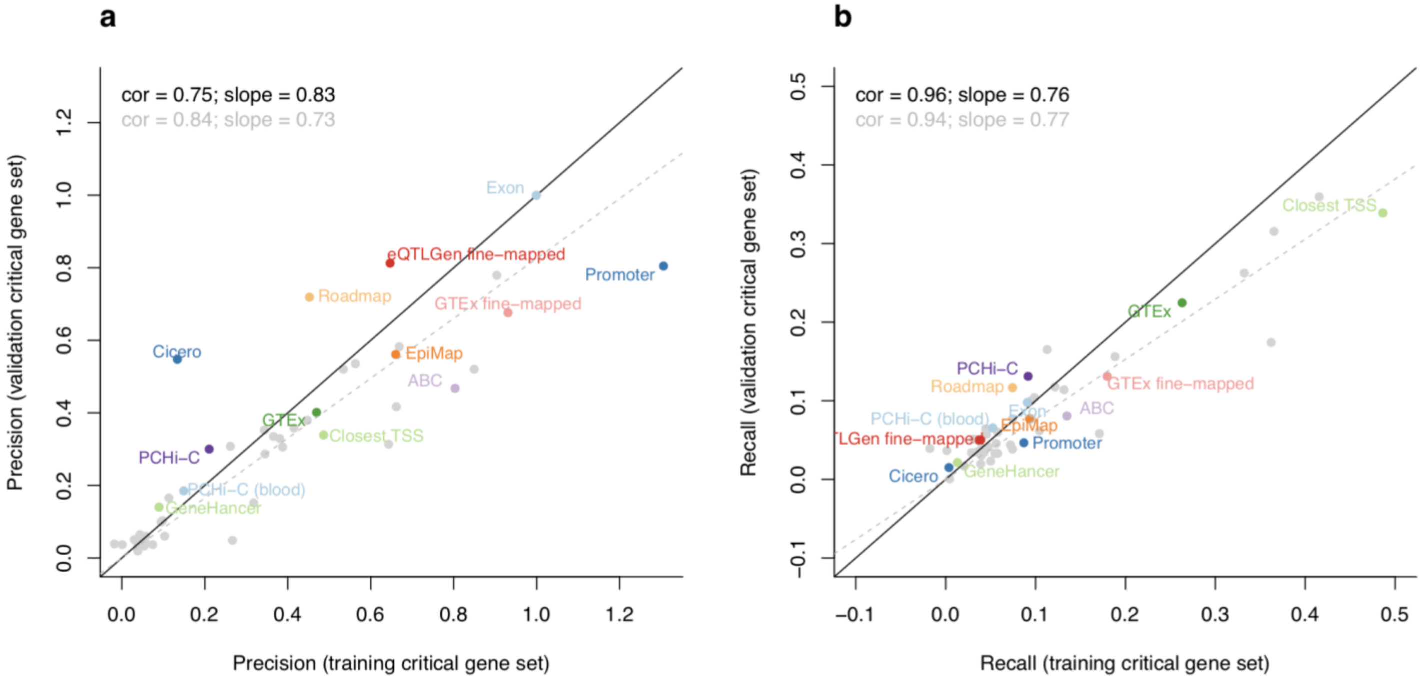


**Supplementary Figure 4: Precision and recall in training and validation critical gene sets.** We compared precision (**a**) and recall (**b**) estimated in our (non-trait-specifics) training critical gene set and (trait-specific) validation critical gene sets. Correlation (cor) and regression coefficient (slope) were computed either using the 13 highlighted independent S2G strategies (see Methods) (results in black), or using all 50 S2G strategies (results in grey). We observed high correlations and slopes for both precision and recall. GTEx: GTEx *cis*-eQTL; GTEx fine-mapped: GTEx fine-mapped *cis*-eQTL; eQTLGen fine-mapped: eQTLGen fine-mapped blood *cis*-eQTL; Roadmap: Roadmap enhancer-gene linking; EpiMap: EpiMap enhancer-gene linking; ABC: Activity-By-Contact

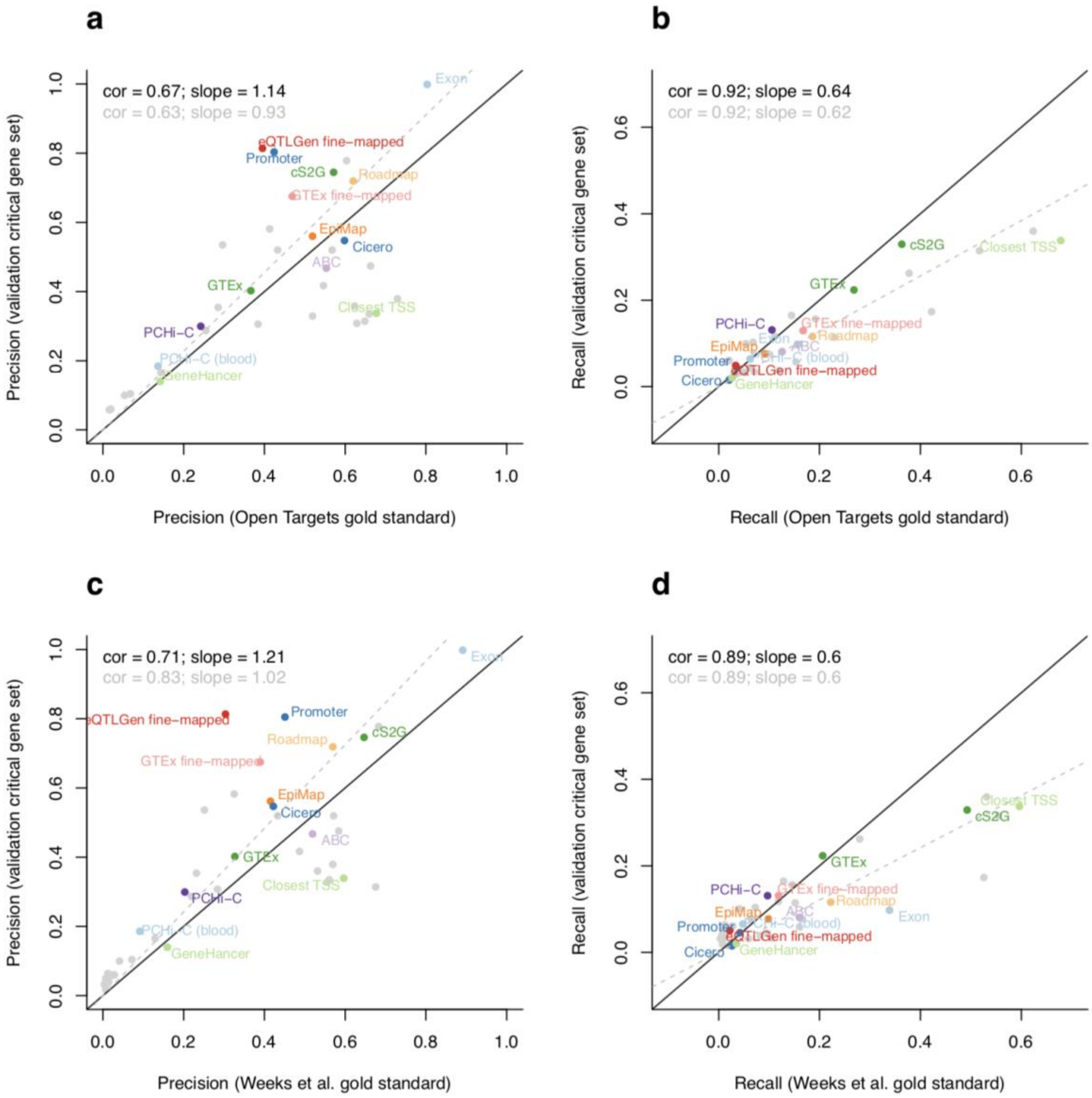


**Supplementary Figure 5: Comparison of precision and recall estimates to independent definitions based on two curated disease-associated lists of SNP-gene pairs.** We compared our estimates of precision and recall to independent definition of precision (i.e. not relying on critical gene sets or polygenic analyses) based on 577 linked sentinel SNP-gene pairs validated with high confidence by Open Targets[11] (**a, b**) and 1,668 linked fine-mapped SNP-gene pairs validated using nearby fine-mapped protein-coding variants[12] (**c, d**). Correlation (cor) and regression coefficient (slope) were computed either using the 13 highlighted independent S2G strategies (see Methods) and the cS2G strategy (results in black), or using all 50 S2G and cS2G strategies (results in grey). We observed high correlations and slopes for both precision and recall. (Note that since recall is the product of $h^2$ coverage and precision, differences in recall basically inherit the differences in precision) Despite the overall concordance, we observed large differences in precision and recall estimates for some S2G strategies (e.g. Exon, Closest TSS), as the curated causal SNP-gene pairs were preferentially ascertained for causal SNPs in which the target gene were the closest one: indeed, we observed an unusually high proportion of pairs involving genes with a short distance (< 10kb) to its closest TSS (57%/67% using both curated lists, vs $h^2$ coverage = 34% for the Closest TSS <10kb S2G strategy). Thus, we caution that curated disease-associated lists of linked SNP-gene pairs may be non-randomly ascertained, highlighting the potential benefits of polygenic analyses for evaluating S2G strategies. GTEx: GTEx *cis*-eQTL; GTEx fine-mapped: GTEx fine-mapped *cis*-eQTL; eQTLGen fine-mapped: eQTLGen fine-mapped blood *cis*-eQTL; Roadmap: Roadmap enhancer-gene linking; EpiMap: EpiMap enhancer-gene linking; ABC: Activity-By-Contact.

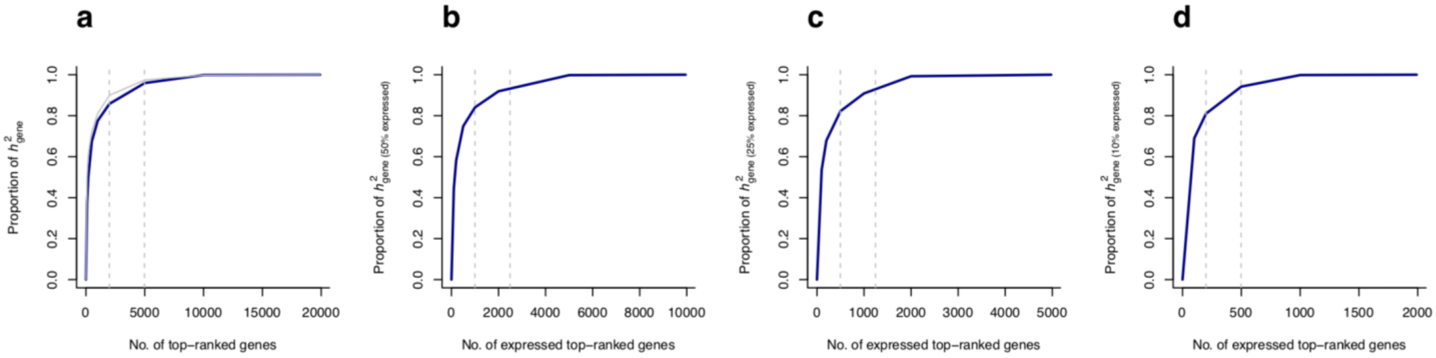


**Supplementary Figure 6: Heritability linked to genes expressed in disease-critical cell-types.** We report the proportion of heritability linked to all genes (**a**; similar than Figure 5a) and linked to genes expressed in disease-critical cell-types (**b,c,d**) explained by genes with the top per-gene $h^2$. We restricted these analyses to 7 of the 16 independent traits that were analyzed in ref.[13] (Chronotype, Diastolic blood pressure, Eczema, Forced Vital Capacity, Mean Platelet Volume, Monocyte Count, and #Children) and plotted the median values. Disease-critical cell-types were selected as in ref.[13] and were glutamatergic for Chronotype, pericyte for Diastolic blood pressure, T-cells for Eczema, smooth muscle for Forced Vital Capacity, megakaryocytes for Mean Platelet Volume, monocytes for Monocyte Count, and GABAergic for #Children. We selected genes expressed in disease-critical cell-types based on the proportion of cells expressed in the cell-types. We selected 50% of the genes with the highest fraction of cells expressed (**b**), 25% of the genes (**c**), and 10% of the genes (**d**). Vertical grey lines indicate 10% and 25% of the genes selected in the analyses and were plotted for comparison purposes. Grey curve in (**a**) indicates results computed on the 16 independent traits (as in Figure 5a) and are similar to the ones computed on the restricted 7 traits. Overall, restricting analyses from Figure 5a to genes expressed in disease-critical cell-types had little impact on the proportion of retained heritability linked to genes explained by the top 10% of retained genes

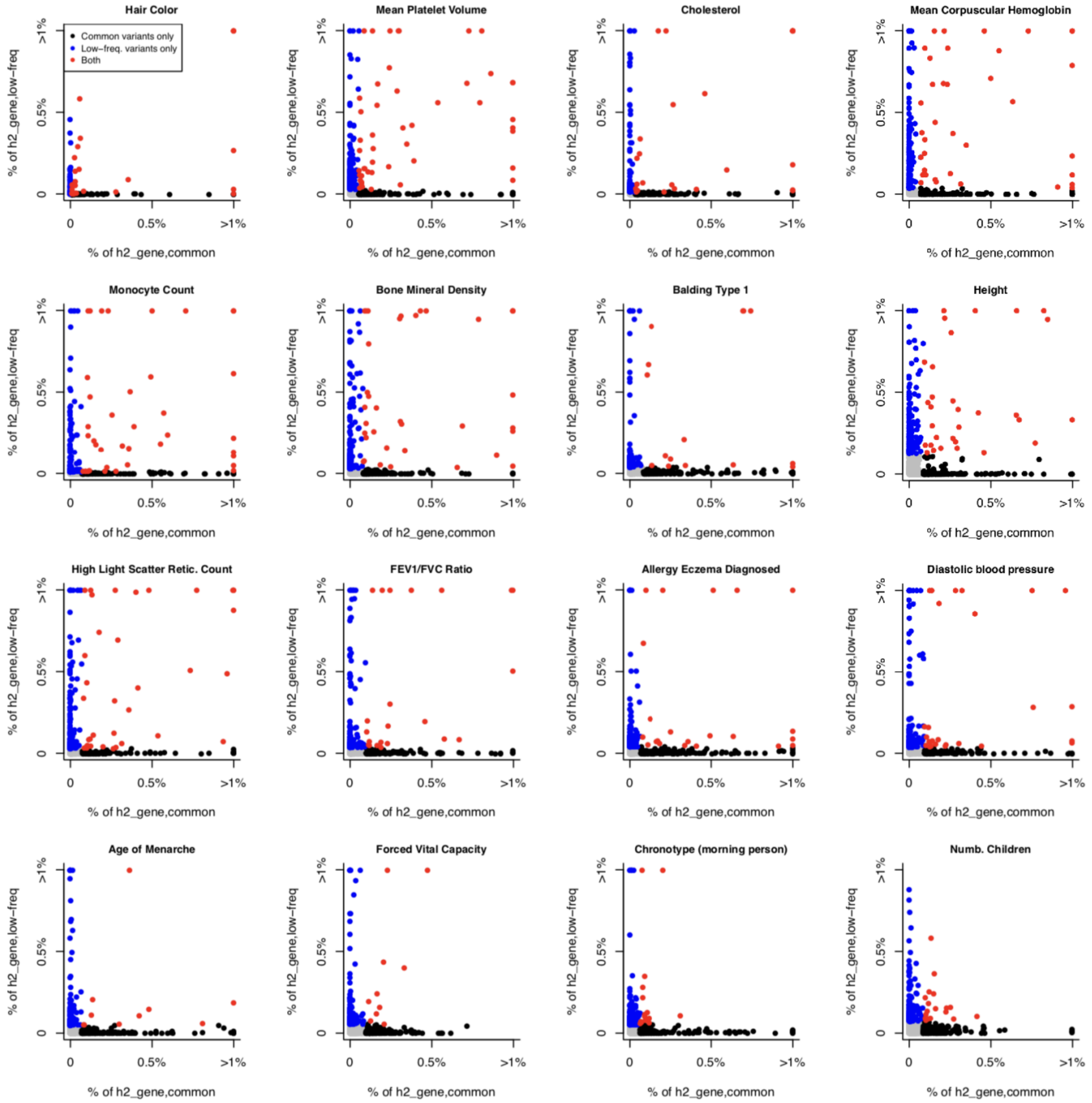


**Supplementary Figure 7: Proportion of common and low-frequency variant heritability linked to genes explained by each individual gene.** We report the proportion of common and low-frequency variant heritability linked to genes ($h^2_{gene,common}$ and $h^2_{gene,low\text{-}freq}$, respectively) explained by each individual gene in 16 independent UK Biobank traits. Genes in the top 200 genes (top 1% of all genes) contributing to both $h^2_{gene,common}$ and $h^2_{gene,low\text{-}freq}$ are denoted in red (median of 26 genes across the 16 traits), genes in the top 200 genes contributing to only $h^2_{gene,common}$ (resp. $h^2_{gene,low\text{-}freq}$) are colored in black (resp. blue) (median of 174 genes each), and remaining genes are colored in grey (median of 19,621 genes, with values close to 0 on both axes). We observe low concordance between per-gene contributions to gene architectures for common vs. low-frequency SNPs.

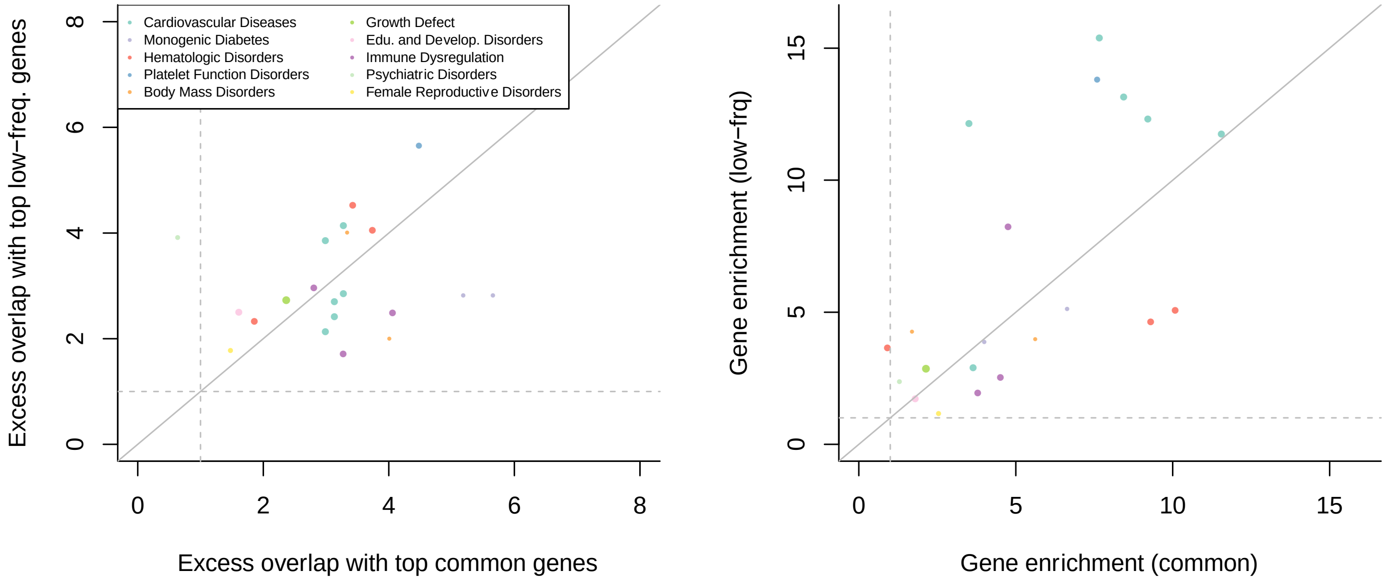


**Supplementary Figure 8: Excess overlap between top genes contributing to common and low-frequency variant heritability linked to genes and disease-specific Mendelian disorder genes.** We report the excess overlap between phenotype-specific Mendelian disorder genes[14] and the top 200 genes contributing to common and low-frequency variant heritability linked to genes (left), and the gene enrichment of disease-specific Mendelian disorder genes (i.e. [heritability linked to Mendelian disorder genes / heritability linked to all genes] / [number of Mendelian disorder genes / total number of genes]) across common and low-frequency variants (right). Each dot represents a disease/trait - Mendelian disorder gene set pair, and is colored by the Mendelian disorder gene set. These two results suggest that both the set of top 200 genes and the per-gene heritability estimates are unlikely to be driven by noisy estimates arising from finite sample size. We restricted analyses to 21 traits analyzed in ref. [14].

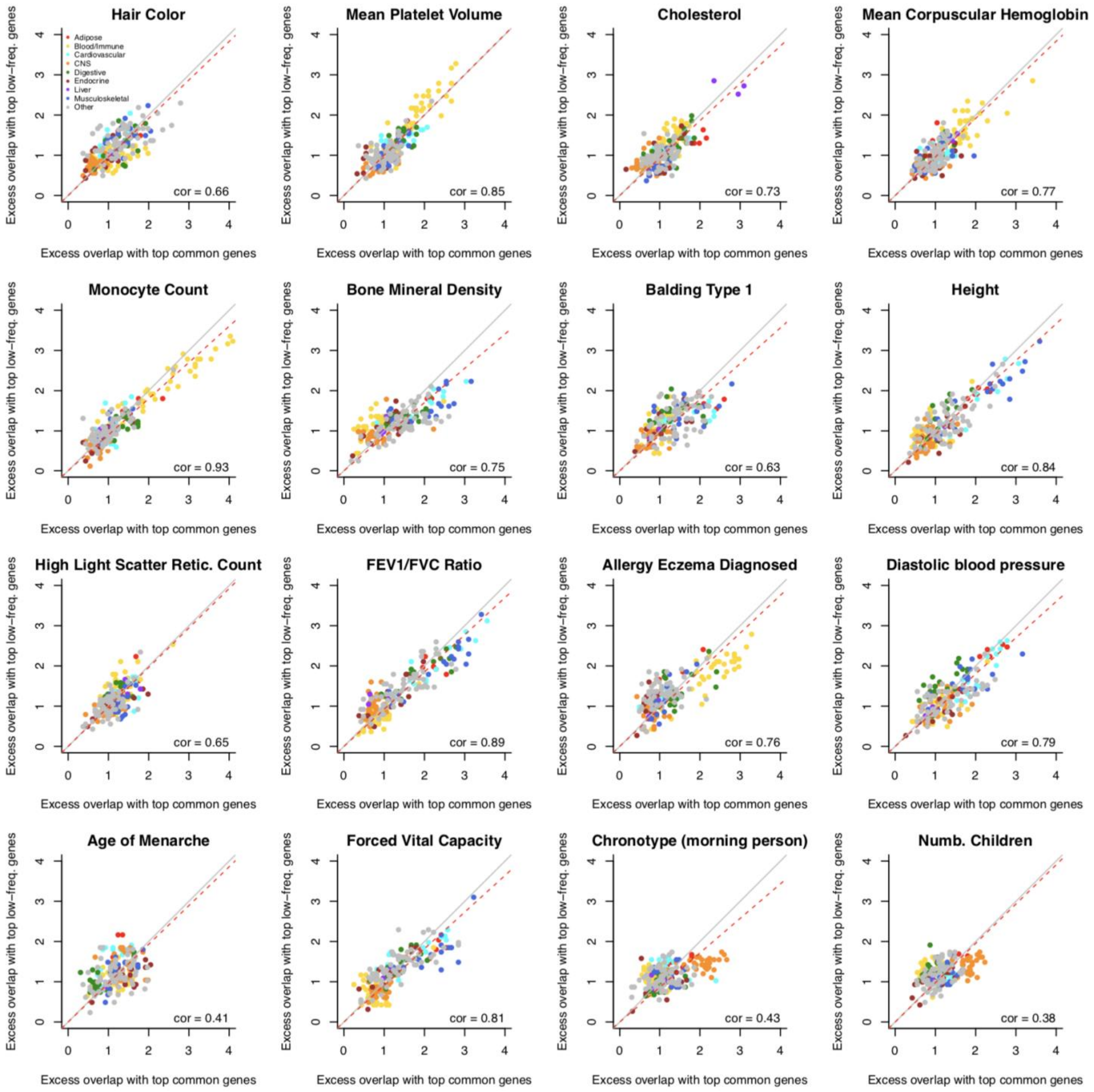


**Supplementary Figure 9: Excess overlap between top genes contributing to common and low-frequency variant heritability linked to genes and differentially expressed gene sets.** We report the excess overlap between 205 differentially expressed gene sets[15] and the top 200 genes contributing to common and low-frequency variants heritability linked to genes across 16 independent UK Biobank traits. Each dot represents a differentially expressed gene set, and is colored by the tissue category. We generally observed excess overlap for disease-critical tissues/cell types. We observed high correlations between excess overlaps for common vs. low-frequency variant architectures, suggesting that common and low-frequency variants architectures are driven by different genes pertaining to similar biological processes.

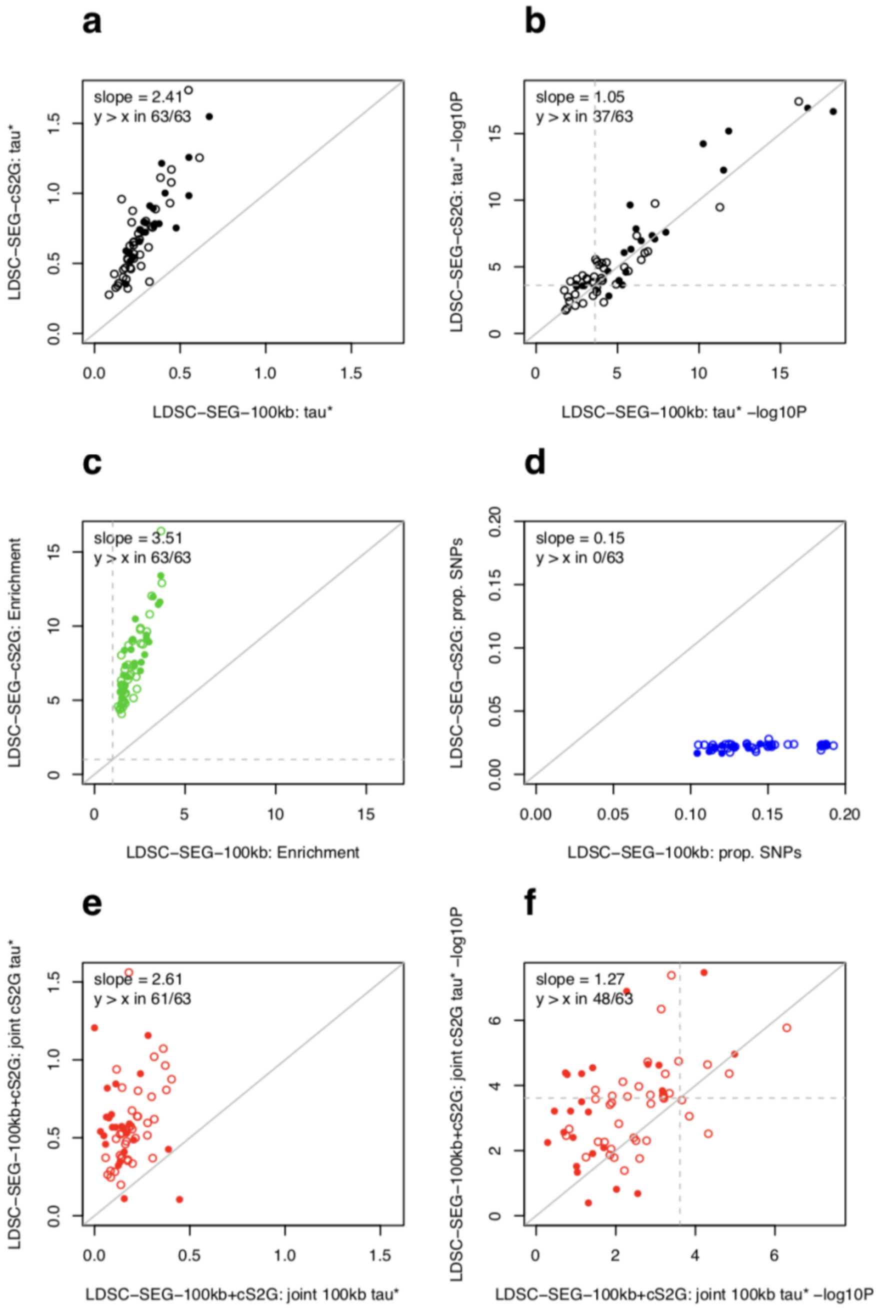


**Supplementary Figure 10: Results of using cS2G vs. ±100kb windows to define SNP annotations in analyses of differentially expressed genes that are enriched for disease heritability.** We compared LDSC-SEG[15] results with the default strategy of linking SNPs to a gene set using a Gene body ± 100kb approach (LDSC-SEG-100kb; *x* axis) to an alternative approach leveraging our cS2G strategy (LDSC-SEG-cS2G; *y* axis). For each of the 63 independent traits, we selected the differentially expressed gene set (out of 205), with the smallest LDSC regression coefficient *P* value (traits with the same gene set selected by LDSC-SEG-100kb and LDSC-SEG-cS2G were represented by a filled dot). We reported the per-standardized-annotation effect sizes $\tau^*$ (**a**), the per-standardized-annotation effect sizes $\tau^*$ *P* value (**b**), the heritability enrichment (**c**), and the proportion of SNPs (**d**) for each 63 trait-gene set pairs. We consistently observed higher $\tau^*$ and heritability enrichment for LDSC-SEG-cS2G (**a,c**). LDSC-SEG-cS2G also produces slightly more significant *P* values than LDSC-SEG-100kb (**b**), despite the fact of losing statistical power by creating SNP annotations with an average of 7 times less SNPs than the default approach (**d**). For 37 out of 63 traits, we obtained more significant *P* values with LDSC-SEG-cS2G; 10 traits have a significant *P* value (<0.05/205) with LDSC-SEG-cS2G but not with LDSC-SEG-100kb, and 5 traits have a significant *P* value with LDSC-SEG-100kb but not with LDSC-SEG-cS2G. In further analyses, we considered a joint model with best gene set SNP annotations from LDSC-SEG-100kb and LDSC-SEG-cS2G (LDSC-SEG-100kb+cS2G). We reported the joint per-standardized-annotation effect sizes $\tau^*$ of the two gene set SNP annotations (**e**), and corresponding *P* value (**f**). We observed that once conditioned to the cS2G gene set SNP annotation, the 100kb gene set SNP annotation was rarely significant (P <0.05/205 in 8/63 traits, against 27/63 with cS2G), suggesting that cS2G captures most of the information in a gene body ± 100kb.

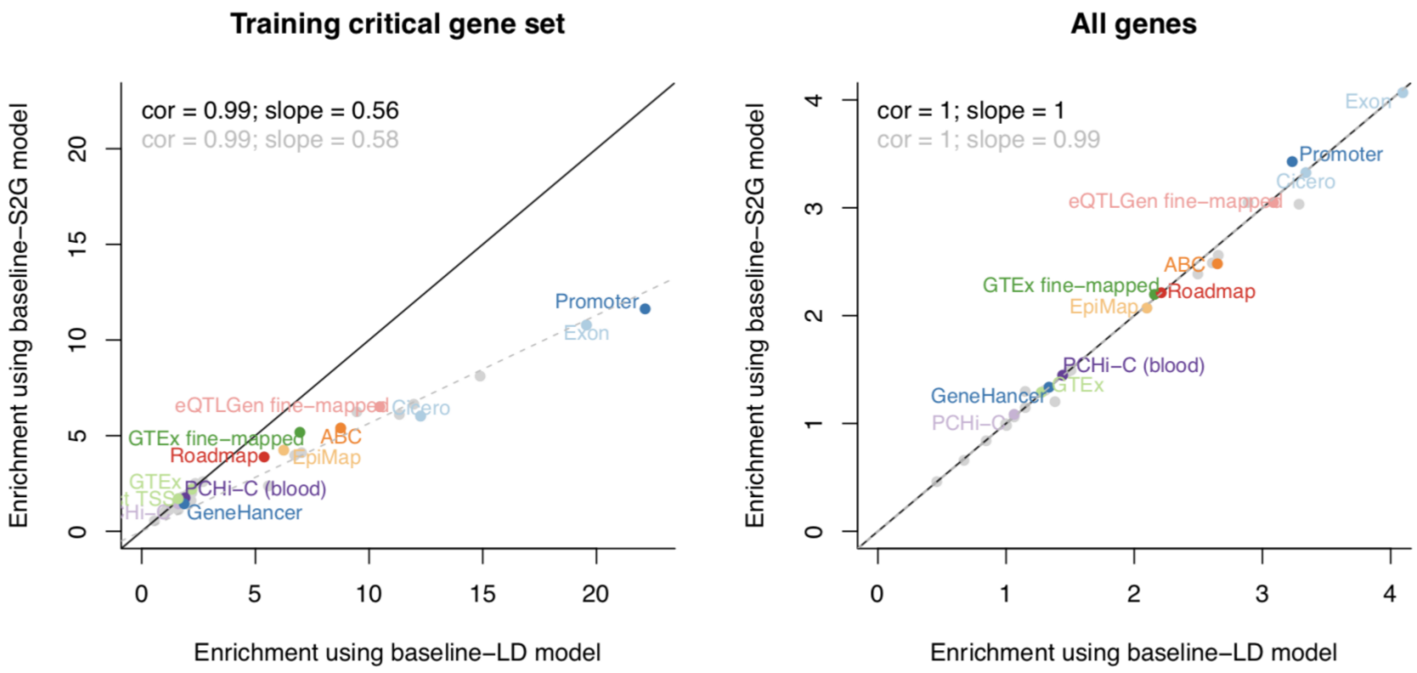


**Supplementary Figure 11: Comparison of heritability enrichment estimates of SNP annotations derived from enriched gene sets using different heritability models.** We report the heritability enrichment of SNP annotations intersecting S2G strategies with constrained genes (left) or all genes (right), estimated by S-LDSC either using the baseline-LD model (and the focal S2G-derived SNP annotation, plus the corresponding S2G-derived SNP annotation for all genes if different) ($x$ axis), or using a model with all baseline-LD SNP annotations and 80 S2G-derived SNP annotations (50 S2G-derived SNP annotations constructed by restricting SNPs linked to genes of the critical gene set, and 30 S2G-derived SNP annotations constructed by restricting SNPs linked to all genes (see Methods); baseline-S2G model) ($y$ axis). Correlation (cor) and regression coefficient (slope) were computed using either the 13 highlighted independent S2G strategies (see Methods) (results in black), or using all 50 S2G strategies (results in grey). We observed that heritability enrichment estimates of SNP annotations intersecting S2G strategies with constrained genes were nearly two times higher when using the baseline-LD model than when using the baseline-S2G model. We thus recommend that future S-LDSC heritability enrichment analyses of gene sets should carefully consider the set of SNP annotations included in the model. We hypothesize that the biases observed under the baseline-LD model are due to tagging effects of unmodeled S2G links; in this case, these biases would not lead to false-positive enriched gene sets. GTEx: GTEx *cis*-eQTL; GTEx fine-mapped: GTEx fine-mapped *cis*-eQTL; eQTLGen fine-mapped: eQTLGen fine-mapped blood *cis*-eQTL; Roadmap: Roadmap enhancer-gene linking; EpiMap: EpiMap enhancer-gene linking; ABC: Activity-By-Contact.

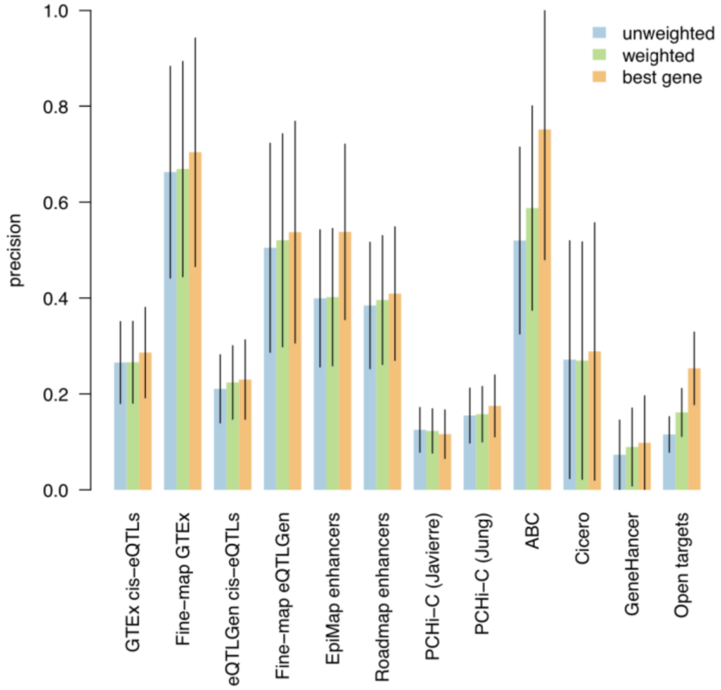


**Supplementary Figure 12: S2G strategy linking each SNP to best gene leads to higher precision than linking SNPs to multiple target genes.** We report the precision of S2G strategies linking SNPs to target genes using three difference approaches forconverting raw linking values into linking scores: by assigning to each gene with non-zero raw linking value the same linking score (unweighted), by assigning to each gene a linking score proportional to its raw linking value (weighted), and by retaining only the gene(s) with the highest linking score (best gene). For most of the S2G strategies the precision was very similar (except for EpiMap, ABC and Open Targets), but the precision was generally highest for the “best gene” strategy.

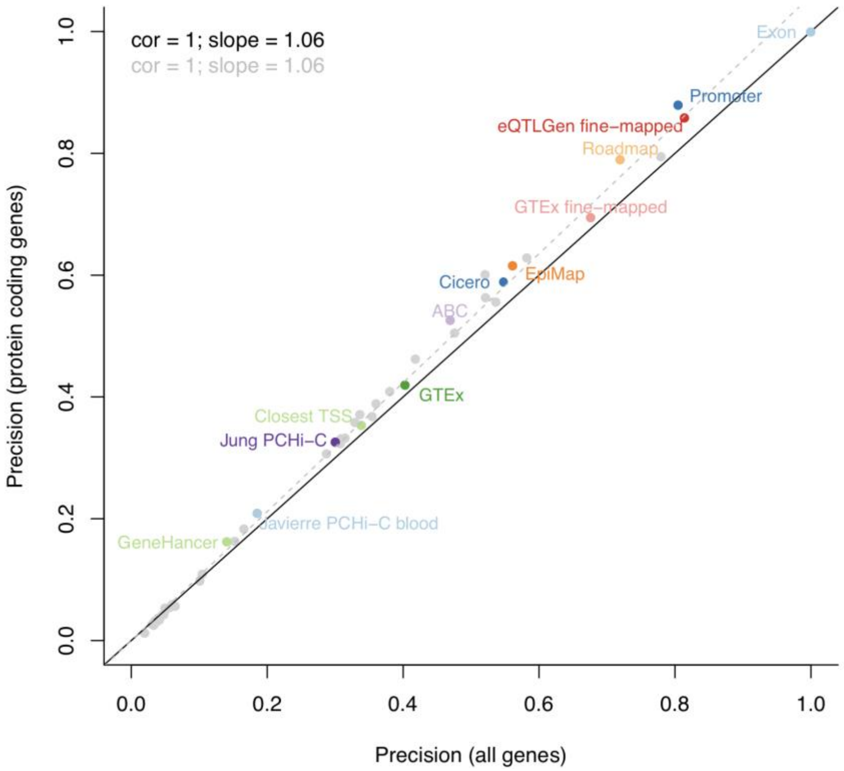


**Supplementary Figure 13: Estimates of precision using 17,871 protein-coding genes instead of 19,995 protein-coding and non-protein-coding genes.** We compared our default estimates of precision using 19,995 protein-coding and non-protein-coding genes to an estimate restricting analyses to 17,871 protein-coding genes. Correlation (cor) and regression coefficient (slope) were computed either using the 13 highlighted independent S2G strategies (see Methods) and the cS2G strategy (results in black), or using all 50 S2G strategies (results in grey). We observed nearly identical estimates. We note that we also used cS2G to estimate that 1.30% of the SNPs linked to non-protein-coding genes (2,124 out of 19,995 genes) explain 1.55 ± 0.18% of heritability, justifying their inclusion even if they are not enriched in heritability ($P$ = 0.71). GTEx: GTEx *cis*-eQTL; GTEx fine-mapped: GTEx fine-mapped *cis*-eQTL; eQTLGen fine-mapped: eQTLGen fine-mapped blood *cis*-eQTL; Roadmap: Roadmap enhancer-gene linking; EpiMap: EpiMap enhancer-gene linking; ABC: Activity-By-Contact.

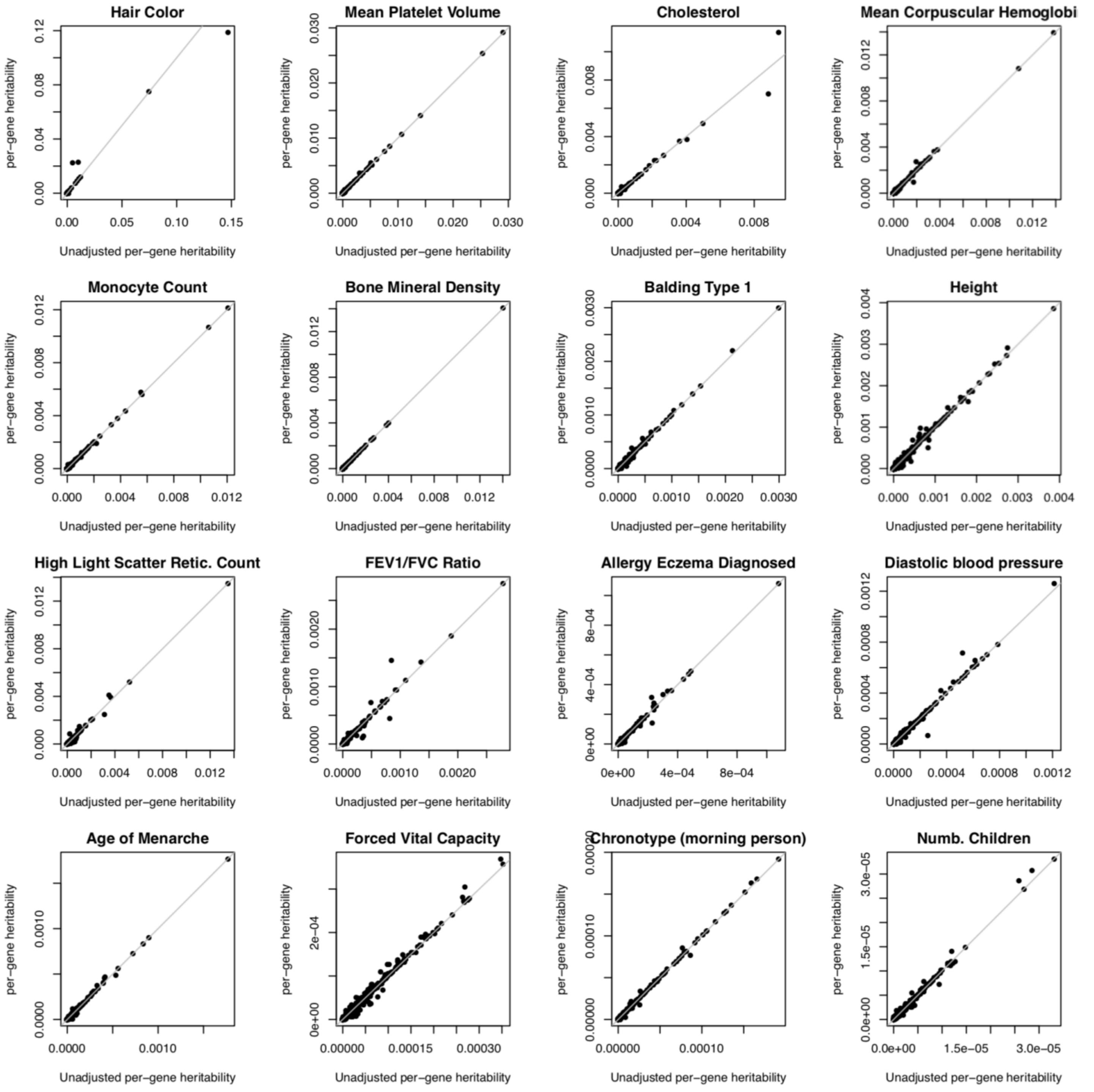


**Supplementary Figure 14: Unadjusted vs. adjusted per-gene heritability estimates.** We report unadjusted per-gene heritability and adjusted per-gene heritability estimates across 16 independent UK Biobank traits. Adjusting per-gene heritability impacted estimates of only a small number of genes.
